# Individuals with Methamphetamine Use Disorder Show Reduced Directed Exploration and Learning Rates Independent of an Aversive Interoceptive State Induction

**DOI:** 10.1101/2024.05.17.24307491

**Authors:** Carter M. Goldman, Toru Takahashi, Claire A. Lavalley, Ning Li, Samuel Taylor, Anne E. Chuning, Rowan Hodson, Jennifer L. Stewart, Robert C. Wilson, Sahib S. Khalsa, Martin P. Paulus, Ryan Smith

## Abstract

Methamphetamine Use Disorder (MUD) is associated with substantially reduced quality of life. Yet, decisions to use persist, due in part to avoidance of anticipated withdrawal states. However, the specific cognitive mechanisms underlying this decision process, and possible modulatory effects of aversive states, remain unclear. Here, 56 individuals with MUD and 58 healthy comparisons (HCs) performed a decision task, both with and without an aversive interoceptive state induction. Computational modeling measured the tendency to test beliefs about uncertain outcomes (directed exploration) and the ability to update beliefs in response to outcomes (learning rates). Compared to HCs, those with MUD exhibited less directed exploration and slower learning rates, but these differences were not affected by the aversive state induction. Follow-up analyses further suggested that reduced exploration in MUD was best explained by greater avoidance of uncertainty on the task, and that trait differences in cognitive reflectiveness might account for these differences in task behavior. These results suggest novel, state-independent computational mechanisms whereby individuals with MUD may have difficulties in testing beliefs about the tolerability of abstinence and in adjusting behavior in response to consequences of continued use.

## Introduction

Methamphetamine Use Disorder (MUD) is characterized by biological, cognitive, and behavioral changes that can be detrimental at both the individual and societal levels. Though outcomes vary widely, common psychological consequences include psychosis, suicidality, hostility, anxiety, depression, and psychomotor dysfunction^1,2^. Despite its growing prevalence worldwide^3^, cognitive mechanisms governing the onset, maintenance, and recurrence of MUD remain unclear.

One means by which MUD may be maintained is through the influence of expected negative outcomes of abstinence and associated withdrawal states, which can motivate avoidance when combined with negative reinforcement processes^4^. In particular, methamphetamine use may attenuate symptoms of depression or somatic anxiety that are brought on or exacerbated by withdrawal. Deficits in interoceptive processing may further contribute to maladaptive behavior, as previous work has shown that individuals with MUD exhibit attenuated neural responses (e.g., insula, anterior cingulate cortex) to aversive somatic states^5^. Countering withdrawal avoidance instead requires that individuals “test out” abstinence as a means of learning whether they are capable of enduring its short-term consequences to improve longer-term quality of life. In computational neuroscience, the abstract structure of this decision problem is captured by so-called “explore-exploit” decision tasks^6,7^. In these tasks, one can either exploit current (limited) knowledge to maximize short-term reward, or one can first test the outcomes of different options (explore) to make better informed choices in the long-term. Importantly, there are different exploratory strategies, which depend on distinct computational processes, and some may be more clinically relevant than others^8–10^. *Directed* exploration (DE), for example, requires keeping track of one’s relative uncertainty about different action outcomes, and then choosing the action for which one has the greatest uncertainty (i.e., as this leads to the most information gain). In contrast, so-called *random* exploration (RE) requires keeping track of one’s total uncertainty across action options, where greater total uncertainty should increase the chance of selecting options that do not currently appear most rewarding (i.e., as one might learn that past experiences were misleading). In the example of withdrawal avoidance mentioned above, DE may be more relevant, as the individual must recognize that they have greater uncertainty about the outcomes of abstinence than about those of continued use. A diminished drive to resolve relative uncertainty could therefore prevent attempts at abstinence and perpetuate the disorder. If it were demonstrated that reduced exploration and sensitivity to uncertainty were present, they could represent possible treatment targets in clinical studies.

Several studies support this in suggesting that iMUDs display lower levels of exploration and altered belief-updating. One study showed that participants with amphetamine use disorder engaged in less information-seeking than healthy controls in a decision-making task that had no cost associated with exploration^11^. This difference may be partly attributable to effects of the drug itself, given that chronic amphetamine use has been shown to deplete intracellular dopamine^12^, and that lower tonic dopamine levels have been linked to lower exploratory behavior in individuals with substance use disorders^13^.

Furthermore, a longitudinal study of iMUDs found that participants who decreased methamphetamine use over a period of six weeks showed higher levels of DE by the end of that period^14^, supporting the idea that methamphetamine use may affect exploratory behavior. In addition to lower levels of exploration, iMUDs have been shown to engage in maladaptive belief-updating, which is often operationalized in terms of altered learning rates within computational models^15–20^. This overall pattern of altered sensitivity to, and learning from, choice outcomes may help explain continued use and high relapse rates despite reduced quality of life^21^.

In this study, we therefore had two main aims. First, we aimed to test whether, compared to healthy comparisons (HCs), iMUDs would show reduced DE and altered learning, as suggested by the literature reviewed above. Second, we sought to test how an aversive interoceptive state (i.e., a somatic anxiety induction) may affect these mechanisms. This allowed us to examine whether physical symptoms typically associated with withdrawal might exacerbate maladaptive decision-making patterns in iMUDs by curtailing further exploration. The latter aim was further motivated by previous work demonstrating increased risk of relapse in substance use disorders under heightened negative affective states^22^, suggesting that sensitivity to current anxiety levels could be an important factor in promoting maladaptive choice within this population. There is also a related body of work indicating relationships between anxiety, exploration, and learning more generally (for a review, see Chou et al.^23^). As the avoidance dynamics driven by these mechanisms are expected to be similar in other substance use disorders (and reflect known maintenance factors in psychopathology more broadly; e.g., anxiety, depression)^24^, these results could also inform future studies with a more transdiagnostic focus.

To accomplish these aims, we fit a computational model with both exploration and learning rate parameters to choice behavior on an established explore-exploit decision task in iMUDs and HCs – both with and without a breathing-based aversive interoceptive state induction. We then tested for both group differences and effects of induced somatic anxiety. We hypothesized that iMUDs would show lower levels of DE than HCs. We also hypothesized that the state anxiety induction would decrease DE in both groups.

As a supplementary aim, we also sought to replicate prior exploratory results demonstrating a relationship between DE and cognitive reflectiveness, and extend this to iMUDs^10^. Cognitive reflectiveness is the tendency to think through a problem before making a decision, instead of simply trusting initial impulses or the first answer that comes to mind^25^. We considered the possibility that lower levels of reflectiveness characteristic of substance use disorder populations (and less reflection on uncertainty in particular) might help to explain reduced DE. This could be of potential clinical relevance, as cognitive reflectiveness has been shown to improve with training^26,27^. Thus, if this hypothesis were confirmed, it would suggest that improving cognitive reflectiveness might promote more adaptive information-seeking in this population. We therefore tested the supplementary hypothesis that iMUDs would differ from HCs in reflectiveness and that this would account for differences in DE within a mediation model.

## Results

To acquire a comprehensive clinical phenotype for iMUDs, participants completed several cognitive and clinical scales (see **Methods**). Compared to HCs, iMUDs showed elevated symptoms of anxiety and depression, higher impulsivity, reduced cognitive performance (working memory), and lower cognitive reflectiveness (see **Table 1**).

**Table 1.**
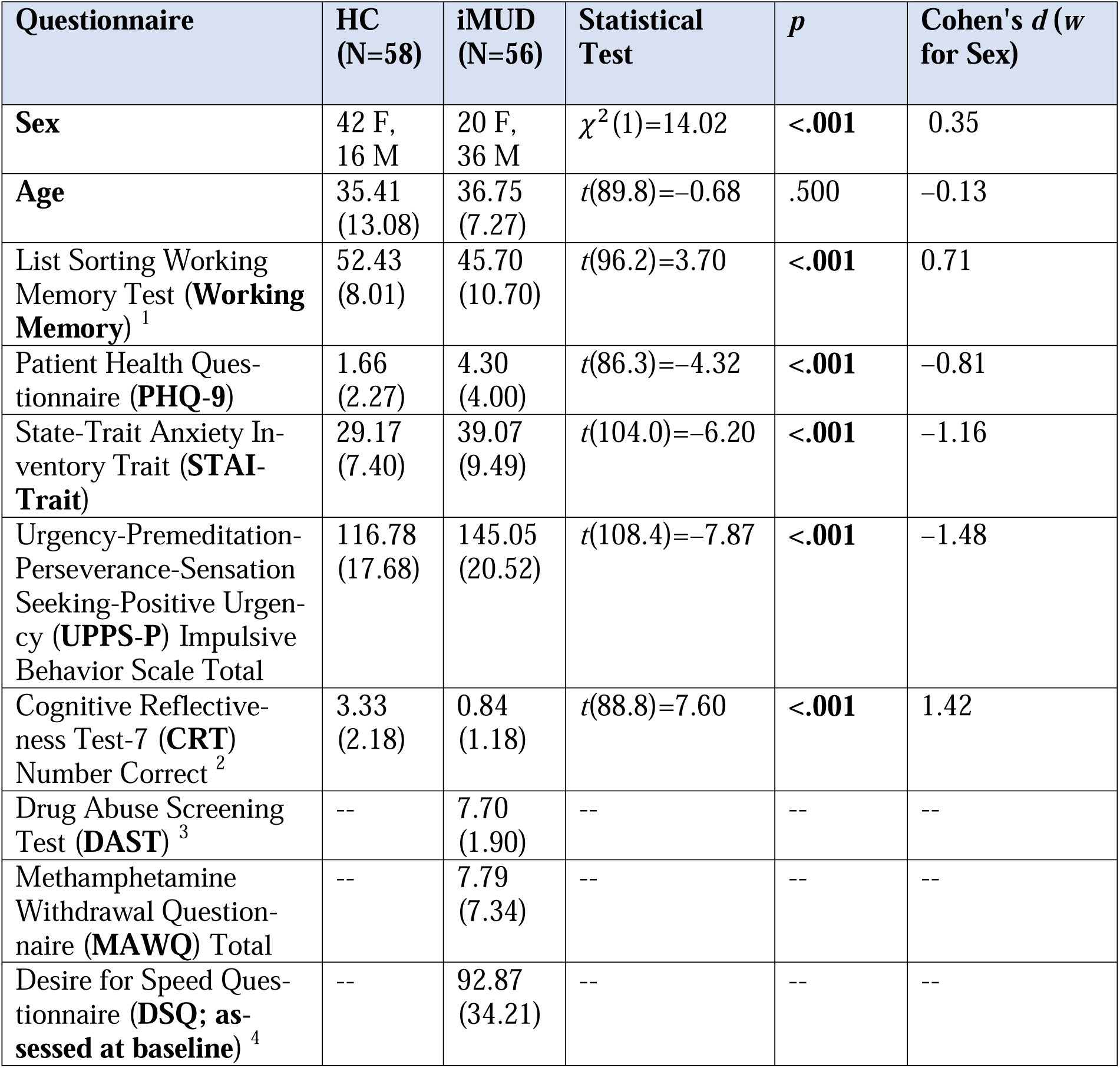
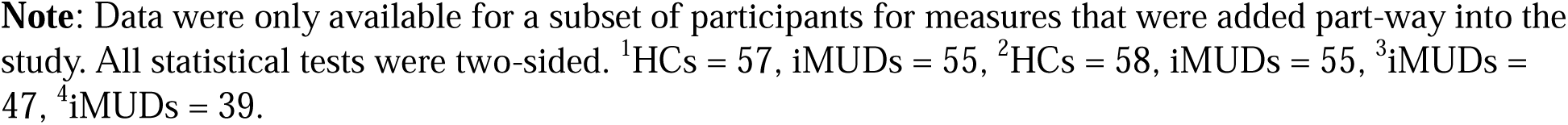
Descriptive Characteristics of HCs and iMUDs.

| Questionnaire | HC<br>(N=58) | iMUD<br>(N=56) | Statistical<br>Test | *p* | Cohen's *d* (*w*<br>for Sex) |
| --- | --- | --- | --- | --- | --- |
| **Sex** | 42 F,<br>16 M | 20 F,<br>36 M | $\chi^2(1)=14.02$ | **<.001** | 0.35 |
| **Age** | 35.41<br>(13.08) | 36.75<br>(7.27) | $t(89.8)=-0.68$ | .500 | -0.13 |
| List Sorting Working<br>Memory Test (**Working**<br>**Memory**)<sup>1</sup> | 52.43<br>(8.01) | 45.70<br>(10.70) | $t(96.2)=3.70$ | **<.001** | 0.71 |
| Patient Health Ques-<br>tionnaire (**PHQ-9**) | 1.66<br>(2.27) | 4.30<br>(4.00) | $t(86.3)=-4.32$ | **<.001** | -0.81 |
| State-Trait Anxiety In-<br>ventory Trait (**STAI-**<br>**Trait**) | 29.17<br>(7.40) | 39.07<br>(9.49) | $t(104.0)=-6.20$ | **<.001** | -1.16 |
| Urgency-Premeditation-Perseverance-Sensation<br>Seeking-Positive Urgen-<br>cy (**UPPS-P**) Impulsive<br>Behavior Scale Total | 116.78<br>(17.68) | 145.05<br>(20.52) | $t(108.4)=-7.87$ | **<.001** | -1.48 |
| Cognitive Reflective-<br>ness Test-7 (**CRT**)<br>Number Correct<sup>2</sup> | 3.33<br>(2.18) | 0.84<br>(1.18) | $t(88.8)=7.60$ | **<.001** | 1.42 |
| Drug Abuse Screening<br>Test (**DAST**)<sup>3</sup> | -- | 7.70<br>(1.90) | -- | -- | -- |
| Methamphetamine<br>Withdrawal Question-<br>naire (**MAWQ**) Total | -- | 7.79<br>(7.34) | -- | -- | -- |
| Desire for Speed Ques-<br>tionnaire (**DSQ; as-**<br>**sessed at baseline**)<sup>4</sup> | -- | 92.87<br>(34.21) | -- | -- | -- |

**Note:** Data were only available for a subset of participants for measures that were added part-way into the study. All statistical tests were two-sided. <sup>1</sup>HCS = 57, iMUDs = 55, <sup>2</sup>HCS = 58, iMUDs = 55, <sup>3</sup>iMUDs = 47, <sup>4</sup>iMUDs = 39.

### The Aversive State Induction Successfully Increased Anxiety During Task Performance Across All Participants

Participants completed the Horizon Task^8^ twice, where one of the runs included a breathing resistance to induce state anxiety. This task requires participants to repeatedly choose between two options to maximize points when given either equal or unequal information about previous reward outcomes from each option (i.e., either two outcomes per side, or one vs. three; based on four initial forced choices). A greater propensity to choose the more uncertain option on subsequent free choices reflects an *information bonus*, while the propensity to choose the less rewarding option reflects *decision noise*. These propensities are moderated by both learning rates from forced-choice outcomes and expected number of future choices (one vs. six; H1 vs. H6 conditions). Namely, information bonus and decision noise should increase from H1 to H6 – where this increase reflects DE and RE, respectively – as exploration could guide future choice in H6 only (see **Methods**).

In a linear mixed-effects model (LME) predicting self-reported anxiety based on group, resistance condition (baseline, task run without resistance, task run with resistance), and their interaction, all effects were significant (*ps*<.001; see **Supplemental Table S2** for full model results), indicating the breathing resistance successfully induced anxiety during the task and this effect was magnified for iMUDs (see **Figure 1**).

**Figure 1.**
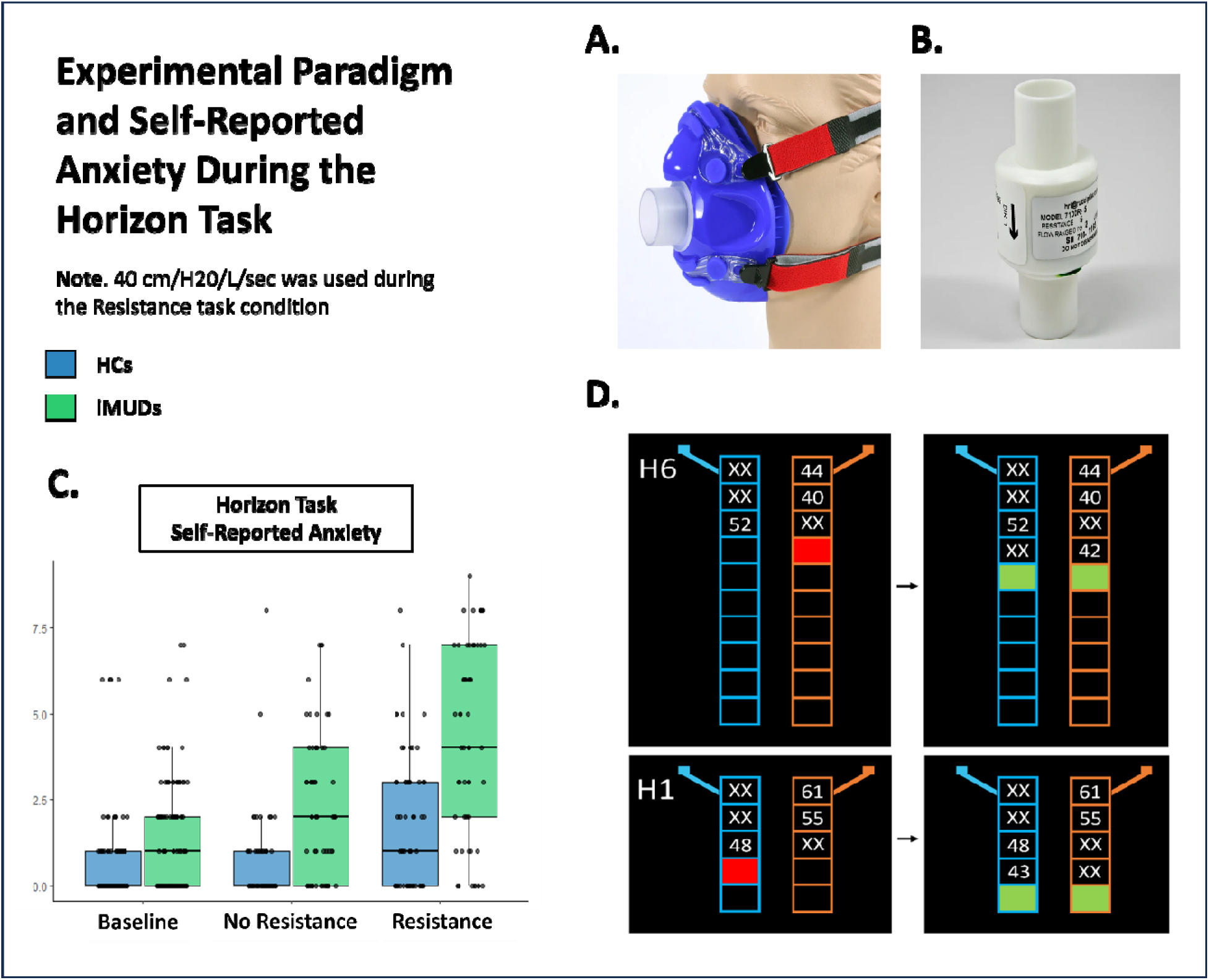
**A)** The silicon mask used during both runs of the Horizon Task. **B)** An example resistor attached to the mask via a plastic tube (not depicted), causing participants to experience resistance during inhalation. A resistance of 40 cmH20/L/sec was used for one of the runs of the Horizon Task to induce somatic anxiety. The other run was completed without breathing resistance. **C)** Boxplots showing the median and quartile values of participants’ self-reported anxiety scores at baseline, during the task run without breathing resistance, and during the task run with breathing resistance. **D)** Horizon Task: Participants first observed outcomes of four forced choices before they were allowed to make either one or six free choices between options to maximize the total number of points received. Games with one or six free choices are referred to as Horizon 1 (H1) and Horizon 6 (H6) games, respectively. The forced choices in each game were either equally informative (two forced choices for each slot machine) or unequally informative (three forced choices for one slot machine and one for the other).

### Individuals with MUD Show Lower Task Performance than Healthy Comparisons

As an initial assessment of task performance, we tested an LME predicting first free-choice accuracy (i.e., choice of the option with the higher average reward value) based on relevant task and experimental conditions. Details are provided in **Supplemental Materials**. In brief, accuracy was higher in H1 than H6 games (as expected), higher in the equal information condition than the unequal information condition, and iMUDs showed lower accuracy than HCs overall. Further, the change in accuracy between H1 and H6 games was less in iMUDs than HCs, consistent with less exploration in iMUDs.

To confirm expected improvements in accuracy over time, and potential modulation of this effect by group or anxiety induction, we tested a subsequent LME predicting accuracy on the six free choices of H6 games based on group, information condition, choice number (1-6), breathing resistance, and the three-way interactions of choice number, group, and breathing resistance, as well as between choice number, group, and information condition (including respective two-way interactions). We observed that accuracy was again higher in HCs (*estimated marginal mean* [*EMM*]=0.81) than iMUDs (EMM=0.69; *contrast[c]*=0.126, *t*(109)=5.42, *p*<.001), higher in the equal than unequal information condition (EMM[equal]=0.77, EMM[unequal]=0.73, *c*=0.040, *t*(2612)=10.73, *p*<.001), and increased as a function of choice number (see **Table 2**). There was also a significant interaction between group and resistance (*F*(1,2612.0)=4.15, *p*=.042), reflecting a numerical increase in accuracy with the breathing resistance in HCs (EMM[Resistance]=0.82, EMM[No Resistance]=0.81, *c*=0.008, *t*(2612)=1.52, *p*=.129) and a numerical decrease in accuracy in iMUDs (EMM[Resistance]=0.68, EMM[No Resistance]=0.69, *c*=0.007, *t*(2612)=-1.36, *p*=.172; see **Figure 2**). All effects remained significant controlling for working memory in a subset of participants for which these scores were available (*p*s<.032).

**Figure 2.**
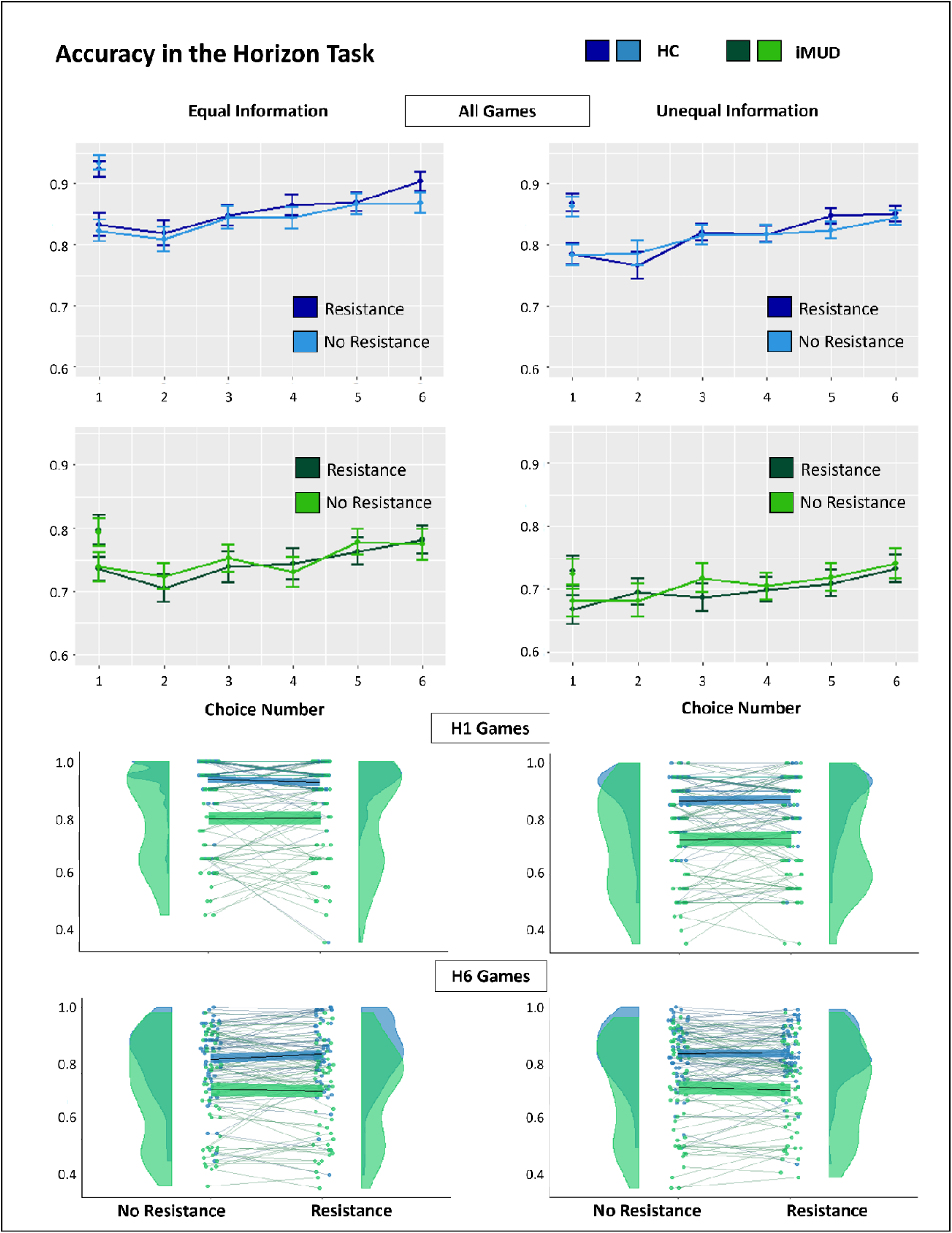
*Top*: H1 and H6 accuracy for each choice by group, resistance, and information condition. Error bars show 95% confidence intervals for accuracy at each choice number. As expected, accuracy was lower in H6 than H1 games for first free choice and improved with further choices in H6 games. The change in first free choice accuracy between H1 and H6 games was also greater in HCs than iMUDs, consistent with greater exploration in HCs. *Bottom*: Accuracy in the H1 and H6 condition for both groups, separated by resistance and information condition. Accuracy was higher in HCs than iMUDs, and greater in equal information games than unequal information games. Breathing resistance increased accuracy for HCs, but decreased accuracy for iMUDs in H6.

**Table 2.** LME Results Predicting Accuracy across H6 Free Choice Trials.

| Outcome Variable | Predictor | Statistical Test | $p$ | $\eta_p^2$ | $b$ [95% CI] |
| --- | --- | --- | --- | --- | --- |
| Accuracy | **Choice Number** | $F(1,2612.0)=124.04$ | **<.001** | 0.04 | 0.012 [0.010, 0.014] |
| | **Group** | $F(1,109.0)=29.35$ | **.001** | 0.21 | -0.063 [-0.086, -0.040] |
| | **Information Condition** | $F(1,2612.0)=114.67$ | **<.001** | 0.04 | -0.020 [-0.024, -0.016] |
| | **Resistance** | $F(1,2612.0)=0.02$ | .899 | 0.00 | 0.000 [-0.004, 0.004] |
| | **Choice Number x Group** | $F(1,2612.0)=1.76$ | .184 | 0.00 | -0.001 [-0.004, 0.001] |
| | **Choice Number x Information Condition** | $F(1,2612.0)=0.13$ | .717 | 0.00 | 0.000 [-0.002, 0.003] |
| | **Group x Information Condition** | $F(1,2612.0)=1.24$ | .265 | 0.00 | -0.002 [-0.006, 0.002] |
| | **Group x Resistance** | $F(1,2612.0)=4.15$ | **.042** | 0.00 | -0.004 [-0.007, 0.000] |
| | **Choice Number x Resistance** | $F(1,2612.0)=1.31$ | .252 | 0.00 | 0.001 [-0.001, 0.003] |
| | **Choice Number x Group x Information Condition** | $F(1,2612.0)=0.00$ | .953 | 0.00 | 0.000 [-0.002, 0.002] |
| | **Choice Number x Group x Resistance** | $F(1,2612.0)=0.46$ | .496 | 0.00 | -0.001 [-0.003, 0.001] |

### Individuals with MUD Show Less Directed Exploration, Random Exploration, and Slower Learning Rates than Healthy Comparisons

The correlations between each of the fitted parameter values were low (rs<.36; see **Supplemental Figure S1**), suggesting that each parameter explained independent aspects of participant behavior.

Further, parameter recoverability was sufficient, indicated by moderate-to-high correlations between parameter values used to simulate behavior and corresponding parameter values estimated from that simulated behavior (.60< *r*s<.89; see **Supplemental Materials Figure S2**).

In our primary computational analyses, LMEs were used to predict model parameter values based on group, resistance level, and their interaction (see **Table 3**). Result showed higher values in HCs for DE (EMM[HC]=6.06, EMM[iMUD]=4.58, *c*=1.48, *t*(109)=2.33, *p*=.021), RE (EMM[HC]=1.67, EMM[iMUD]=1.04, *c*=0.63, *t*(109)=1.98, *p*=.050), *α*_0_ (initial learning rate), and *α*_∞_ (asymptotic learning rate; i.e., the value to which a participant’s learning rate would theoretically converge if the game were played indefinitely; EMM[HC]=0.30, EMM[iMUD]=0.22, *c*=0.077, *t*(118.3)=3.06, *p*=.003). Notably, the group difference in RE was no longer significant after two potential outliers were removed using a Grubb’s test (*F*(1,110.0)=1.59, *p*=.209, *η_p_*^2^ =0.01), indicating high values that fell sufficiently outside the overall sample distribution. Because *α*_0_ values showed a bimodal distribution across participants (see **Figure 3**), we instead performed a k-means clustering analysis and divided participants into those with high and low values, and then used cluster membership as a categorical outcome variable in logistic mixed regressions in place of LMEs. In these regressions predicting *α*_0_, we observed a significant group difference (proportion in high-value cluster: HCs=0.67; iMUDs=0.32; see **Table 3** for statistical results). There was no main effect of breathing resistance or interaction with group for any parameter.

**Figure 3.**
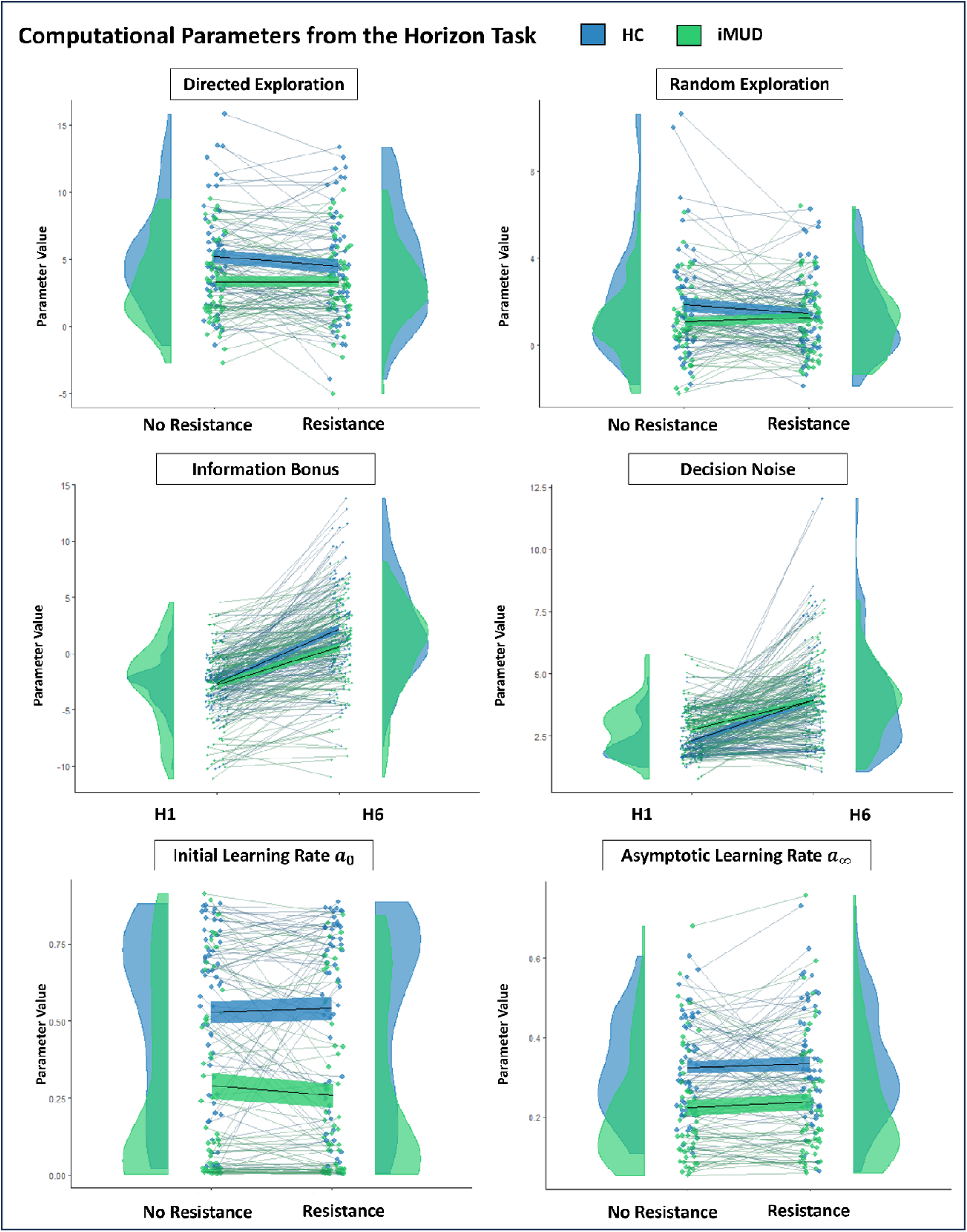
Plots of participants’ computational parameters for the Horizon Task, where each participants’ parameter values are connected by a line. The black line and surrounding ribbon mark the mean and standard error, respectively, of parameter values for each group.

**Table 3.** Results Testing Effects of Group and Resistance on Primary Computational Measures.

| Outcome Variable | Predictor | Statistical Test | $p$ | $\eta_p^2$ | $b$ [95% CI] |
| --- | --- | --- | --- | --- | --- |
| DE | Group | $F(1,109.0)=5.45$ | **.021** | 0.05 | -0.742 [-1.365, -0.119] |
| | Resistance | $F(1,112.0)=1.63$ | .204 | 0.01 | -0.187 [-0.480, 0.105] |
| | Group x Resistance | $F(1,112.0)=1.56$ | .214 | 0.01 | 0.186 [-0.106, 0.479] |
| RE | Group | $F(1,109.0)=3.93$ | **.050** | 0.03 | -0.316 [-0.628, -0.004] |
| | Resistance | $F(1,112.0)=0.32$ | .575 | <0.01 | -0.060 [-0.277, 0.158] |
| | Group x Resistance | $F(1,112.0)=1.85$ | .177 | 0.02 | 0.151 [-0.067, 0.369] |
| Initial Learning Rate | Group | $\chi^2(1)=11.02$ | **<.001** | -- | -1.800 [-2.859, -0.742] |
| | Resistance | $\chi^2(1)=0.15$ | .695 | -- | -0.080 [-0.466, 0.306] |
| | Group x Resistance | $\chi^2(1)=0.16$ | .685 | -- | -0.080 [-0.466, 0.306] |
| Asymptotic Learning Rate | Group | $F(1,118.4)=9.42$ | **.003** | 0.07 | -0.039 [-0.063, -0.014] |
| | Resistance | $F(1,111.6)=0.88$ | .351 | <0.01 | 0.006 [-0.007, 0.019] |
| | Group x Resistance | $F(1,112.0)=0.18$ | .674 | <0.01 | 0.003 [-0.010, 0.016] |

Complementary Bayesian models predicting each learning rate based on group, breathing resistance, and their interaction allowed incorporation of information regarding posterior variances around each parameter estimate. These analyses provided further evidence for lower values of *α*_0_ (*b*=-0.15, BCI=[-0.22, -0.09], BF=51.09) and *α*_∞_ (*b*=-0.06, BCI=[-0.09, -0.04], BF=14.48) in iMUDs. Notably, however, when *α*_0_ was included as a covariate in the model predicting *α*_∞_, we did not find evidence for group differences (*b*=-0.03, BCI=[-0.06, -0.01], BF=.04).

When working memory was included as an additional covariate in the subset of participants with available data, effects of group in the LMEs above remained significant for all parameters (*p*s<.034), except in relation to DE, which was no longer significant (*F*(1,103.0)=3.09, *p=.*082, *η_p_*^2^ =0.03, *b=*–0.614, CI=[-1.1.298, 0.071]; see **Supplemental Table S7**). This suggested poor working memory might contribute to reduced DE in iMUDs. However, working memory was not itself a significant predictor in any model (*ps>*.174).

To better interpret these group differences, effects on parameters in H1 and H6 were then examined separately. In an LME predicting the information bonus parameter by horizon, group, and their interaction, the interaction was significant (*F*(1,340.0)=12.60, *p*<.001, *η_p_*^2^ =0.04, *b*=-0.383, CI=[-0.595, -0.172]), reflecting the group difference in DE described above (see **Figure 3**). Post-hoc contrasts showed that HCs had higher information bonus values than iMUDs in H6 (HCs–iMUDs=1.66, *t*(139.1)=2.63, *p=*.010), but not in H1 (HCs–iMUDs=0.13, *t*(139.1)=0.20, *p=*.843). Notably, this interaction remained significant when accounting for working memory (*F*(1,325.0)=10.72, *p<.*001, *η_p_*^2^ =0.03, *b*=-0.360, CI=[-0.575, -0.144]), supporting an independent group difference in DE. Complementary Bayesian analyses incorporating the posterior variance around information bonus estimates provided further evidence for this interaction between group and horizon (*b*=-0.38, BCI=[-0.63,-0.14], BF=38.78).

In an analogous LME predicting decision noise based on horizon, group, and their interaction, the interaction was not significant (*F*(1,340.0)=3.68, *p=.*056, 1^2^ =0.01, *b*=-0.117, CI=[-0.237, 0.003]).

However, this interaction became significant when accounting for working memory in the subset of participants with available data (*F*(1,325.0)=4.13, *p=.*043, *1*^2^ =0.01, *b*=–0.129, CI=[-0.253, -0.005]). Post-hoc contrasts here suggested that group differences in RE were driven by (numerically) greater decision noise in iMUDs than HCs in H1 (HC–iMUD=–.40, *t*(212.9)= –1.81, *p=*.072), with no difference in H6 (HC–iMUD=0.12, *t*(212.9)=0.53, *p=*.598; see **Figure 3**). However, complementary Bayesian analyses did not provide support for this interaction (*b*=-0.02, BCI=[-0.15,0.11], BF=0.18).

### Individuals with MUD Showed Greater Avoidance of Uncertainty

As a complementary test of uncertainty avoidance, we calculated the probability of choosing the option with the greater observed mean reward when it was also the option with greater uncertainty (i.e., only one forced choice outcome shown) on the first free choice for H1 and H6 games separately. Here, we reasoned that greater uncertainty avoidance would be reflected by fewer choices of the uncertain option despite the greater observed reward. To evaluate this, we tested an LME predicting this probability for H6 games based on group and resistance condition. The corresponding probability for H1 games was also included in the model to account for any baseline tendency to approach or avoid uncertainty when exploration would not be helpful. This LME revealed a main effect of group, such that iMUDs had a lower tendency to choose the option with a greater mean reward value when this option had greater uncertainty (EMM[HC]=0.82, EMM[iMUD]=0.74, *F*(1,117.2)=6.47, *p*=.012, *b*=–0.039, CI=[-0.069, -0.009]). However, the effect of group became nonsignificant when working memory was included in the model (*F*(1,108.2)=3.80, *p*=.054, *b*=–0.032, CI=[-0.064, 0.001]). An analogous LME was run to test if negative outcome avoidance differed between groups. This model instead predicted the probability of choosing the option with the greater observed mean reward when it was also the option with lower uncertainty in H6 games based on group and resistance condition, while again controlling for baseline tendencies in H1. Here, we reasoned that those with greater negative outcome avoidance would more often choose the option with the higher observed value, despite the greater information gain afforded by the other option. In this case, there was no effect of group (EMM[HC]=0.67, EMM[iMUD]=0.66, *F*(1,112.3)=0.02, *p*=.895, *b*=–0.002, CI=[-0.039, 0.034]), supporting the idea that HCs and iMUDs primarily differed in their avoidance of uncertainty rather than avoidance of negative outcomes.

### No Relationships were Observed Between Computational Parameter Values and Somatic Anxiety, Substance Use Symptoms, or Measures of Psychopathology

To determine if somatic anxiety related to the patterns of behavior captured by the model, we estimated LMEs predicting each computational parameter value based on self-reported anxiety during the task. This tested whether those with greater state anxiety also had greater changes in behavior. Among other covariates, effects of group and resistance condition were also included, as well as the interaction between resistance condition and self-reported anxiety. This interaction captured the possibility that those who were more sensitive to the somatic manipulation (i.e., greater increases in anxiety) also showed greater changes in exploratory behavior. In these models, we did not observe any effects of self-reported anxiety (*p*s>.266) or its interaction with resistance condition (*p*s>.117). Thus, we did not find evidence that individual differences in sensitivity to the somatic manipulation were related to exploratory tendencies or other patterns of behavior in the task.

Follow-up LMEs were run predicting model parameters based on substance use symptoms in iMUDs (i.e., DAST, DSQ, MAWQ; tested separately), breathing resistance, and their interaction. In all models, we observed no significant main effects of substance use symptoms nor interaction effects (see **Supplemental Tables S8-S10**). However, in the models predicting *α*_∞_ based on MAWQ and DAST (separately), we observed significant effects of breathing resistance when working memory was included as a covariate (see **Supplemental Tables S8** and **S9**). Considering the absence of an effect of breathing resistance in other computational analyses, and that these results were not hypothesized, we simply note them here for the interested reader and for purposes of future hypothesis generation.

LMEs testing potential parameter differences within iMUDs based on comorbid psychopathology (present/absent), continuous measures of psychopathology (PHQ-9, UPPS-P Total, and STAI-Trait Scores; tested separately), medication status (medicated/unmedicated), time since last methamphetamine use, and time since starting treatment did not show any significant effects (*ps* >.153).

### Greater Random Exploration and Faster Learning Rates Each Improved Performance

To evaluate the theoretical significance of observed group differences, we tested whether some values for each model parameter might be considered more optimal than others with respect to task performance after the first free choice in H6. In brief, LMEs revealed that higher values of *α*_0_ and *α*_∞_ each predicted greater accuracy in equal information games, though only higher values of *α*_0_ predicted greater accuracy in unequal information games (*p*=.010). Higher values of RE predicted steeper improvements in accuracy over time for equal information games, while higher values of *α*_0_ predicted steeper improvements in unequal information games (*p*s<.041). We did not observe a relationship between DE and accuracy in unequal information games (*F*(1,758.2)=1.72, *p*=.191, *η_p_*^2^ <.01, *b*=0.002 [-0.001, 0.005]); nor did we observe that DE related to greater improvements over time (although the latter result was trending in the expected direction; *F*(1,977.1)=3.27, *p*=.071, *η_p_*^2^ <.01, *b*=0.001 [-0.000, 0.002]; see **Methods** and **Supplemental Tables S11** and **S12**).

### Directed Exploration and Learning Rates were Predicted by Cognitive Reflectiveness

As a secondary aim, we sought to replicate and extend our prior results linking exploration to cognitive reflection^10^. To do so, we tested LMEs predicting model parameters based on Cognitive Reflection Test (CRT) scores, accounting for potential effects of group and resistance. Across all participants, CRT score significantly predicted DE (*F*(1,107.0)=3.95, *p=.*050, *η_p_*^2^ =0.04, *b=*0.344, CI=[0.001, 0.683]) and *α*_0_ (*χ*^2^(1)=7.04, *p=.*008, *b=*0.699, CI=[0.182, 1.215]), but not *α*_∞_ (*F*(1,109.7)=0.13, *p=.*719, *η_p_*^2^ <0.01, *b=*0.002, CI=[-0.011, 0.015]) or RE (*F*(1,107.0)=0.06, *p=.*811, *η_p_*^2^ <0.01, *b=*0.021, CI=[-0.152, 0.195]). Notably, when additionally controlling for working memory, CRT remained a significant predictor of *α*_0_ (*χ*^2^(1)=7.24, *p=.*007, *b=*0.683, CI= [0.173, 1.104]) but was no longer a significant predictor of DE (*F*(1,101.0)=3.81, *p*=.054, *η_p_*^2^ =0.04, *b*=0.347, CI=[-0.001, 0.696].

For analogous models restricted to the iMUD sample, CRT score did not significantly predict DE, RE, or *α*_∞_ (*p*s>.189); nor was it a significant predictor of *α*_0_ (*F*(1,51.0)=3.68, *p=.*061, *η_p_*^2^ =0.07, *b=*0.066, CI=[-0.003, 0.134]). There were also no observed effects of resistance (*p*s>.191). Note that, unlike in the full sample, in iMUDs alone the distribution of *α*_0_ values was sufficiently normal to use an LME in place of logistic regression.

### Cognitive Reflectiveness Accounts for Group Differences in Directed Exploration and Learning Rates

The group difference in computational parameters observed above motivated us to test mediation models in which lower values of DE and *α*_0_ in iMUDs might be explained by lower reflectiveness (CRT scores). These mediation models included group as the predictor variable and either DE or *α*_0_ as the outcome variable, with CRT score as the potential mediator. In the mediation model predicting DE (*total effect c*=-0.73, *p*=.024, CI=[-1.364, -0.10]), we observed a significant indirect effect (*ab*=-0.50, *p*=.045, CI=[-1.016, -0.01]), and a non-significant direct effect (*c*=-0.24, *p*=.567, CI=[-0.235, 0.54]), supporting CRT as a mediator. In the mediation model for *α*_0_ (*total effect c*=-0.15, p<.001, CI=[-.200, -0.09]), we also observed a significant indirect effect (*ab*=-0.062, *p*=.004, CI=[-0.109, -0.02]), and a non-significant direct effect (*c*=-0.083, *p*=.014, CI=[-0.150, -0.02]), again supporting CRT as a mediator. This suggested that lower levels of reflectiveness on uncertainty in iMUDs may contribute to the lower levels of DE and slower learning rates observed in this group.

## Discussion

In the present study, we compared how treatment-seeking (currently abstinent) individuals with Methamphetamine Use Disorder (iMUDs) and healthy comparisons (HCs) differed in information-seeking and learning under uncertainty, both with and without a somatic anxiety induction. This allowed us to distinguish the causal effect of state anxiety from potential effects of other factors linked to psychopathology. As expected, we found that HCs outperformed iMUDs on the task. Computational modeling revealed that iMUDs had lower values of directed exploration (DE), random exploration (RE), initial learning rate (*α*_0_), and asymptotic learning rate (*α*_∞_; controlling for *α*_0_), while these parameters themselves were only weakly correlated.

The differences observed in DE and RE support previous research finding that individuals with substance use problems exhibit reduced exploration^28,29^. Importantly, however, unlike several previous studies, the Horizon Task allowed us to distinguish directed from random strategies, where measures of DE and RE in this task are also sensitive to beneficial vs. suboptimal engagement in exploratory behavior (i.e., with vs. without future choices that could benefit from information gain). Here, DE differences in iMUDs appeared to reflect an attenuated ability to increase exploration when it was beneficial (i.e., in games with a longer horizon) and a greater motivation to avoid uncertain options. In contrast, differences in RE were attributable to less reward-sensitive choices when this was not beneficial (i.e., in games with a shorter horizon). However, group differences in RE were no longer significant after potential outliers were removed and were not supported by complementary Bayesian analyses; these results should therefore be treated with caution. Overall, these findings offer insights into more specific cognitive mechanisms that might contribute to maladaptive choice and withdrawal avoidance.

The differences observed in initial and asymptotic learning rates suggest iMUDs update their beliefs less after observing each new outcome, representing a possible overconfidence in prior beliefs. This could be taken to reflect a greater expectation that mean reward values would remain stable.

Alternatively, it could indicate the belief that each observed outcome would be less informative (i.e., more noisy, less trustworthy) with respect to the true value of the underlying reward mean^30^. In real-world contexts, slower learning rates can prevent individuals from changing their behavior, despite experiencing harmful consequences. However, it remains to be shown whether such results generalize to learning in daily life in this population.

Contrary to our hypothesis, somatic anxiety induction did not affect any computational measure. Here, it is notable that, while previous research has shown negative correlations between trait anxiety and DE^9,10^, to our knowledge, this is the first study to causally manipulate somatic state anxiety and differentiate its influence from trait factors in iMUDs. While this could indicate that state anxiety does not account for differences in information-seeking, it is possible that the resistance level, which was chosen to maintain tolerability, did not induce sufficiently high anxiety. Self-reported anxiety was higher for the task run with resistance (Mean=3.05) than without resistance (Mean=1.46), and this effect was greater for iMUDs than HCs, but anxiety scores were still well below the maximum score of 10. Future work might therefore aim to induce higher levels of somatic anxiety in a feasible manner and reassess its potential effects. Incorporating trait measures of interoceptive awareness could also help clarify whether the relationship between somatic anxiety and decision-making is altered in those with higher sensitivity. On the other hand, our results do appear consist with some prior work showing no change in model-based planning after anxiety induction^31^. To the extent that DE depends on model-based processes, these results could point in a similar direction.

In line with our secondary aim, results also successfully replicated prior findings^10^ linking cognitive reflectiveness to DE, and also showed a novel association with initial learning rates. Mediation analyses also suggested group differences in DE and initial learning rate might be accounted for by differences in cognitive reflectiveness. This suggests that lower reflectiveness may reduce adaptive information-seeking in iMUDs and interfere with learning in uncertain environments. It should be stressed, however, that these analyses on trait reflectiveness were cross-sectional and do not support causal inference. They simply highlight shared explanatory variance between these measures that could offer additional insights. We also note that the relationship between cognitive reflectiveness and model parameters was not observed in iMUDs alone. This could be due to insufficient sample size, the lower values and restricted range of reflectiveness scores in iMUDs, or perhaps a mechanism whereby substance use decouples these variables. Future work should examine whether improving reflectiveness could promote more adaptive information seeking and learning, and whether this might be clinically beneficial. This possibility is supported by previous work showing that cognitive reflectiveness can be improved with training^26,27^.

It is important to consider limitations of the present study when interpreting these results. First, our sample size was only moderate and unable to reliably detect small effect sizes. We also could not determine whether observed group differences represent a pre-existing vulnerability factor or a consequence of methamphetamine use. No relationships were found with length of abstinence, days since starting treatment, or medication status, perhaps suggesting that group differences were better explained by pre-existing factors or were insensitive to recovery; however, longer recovery times will need to be examined. The presence of other comorbid affective and substance use disorders also did not appear to account for any results.

With these limitations in mind, we found that individuals with methamphetamine use disorder exhibited lower levels of exploration and reduced learning rates when making decisions under uncertainty. Contrary to expectation, we did not observe an effect of aversive interoceptive state induction (and resulting increases in somatic anxiety) on model parameters or other behavioral metrics, suggesting trait factors may be of more central importance. Overall, these results highlight directed exploration and learning rates, and underlying uncertainty estimation processes, as possible mechanisms of maladaptive choice in individuals with methamphetamine use disorder and could point to novel treatment targets that could be tested in future work.

## Methods

### Participants

Participants included 56 inpatient treatment-seeking iMUDs and 58 HCs. Individuals with MUD were currently abstinent (mean time since methamphetamine use=50.04 days, mean time since starting treatment=34.47 days) and recruited from two recovery homes in the Tulsa, Oklahoma area: (1) GRAND and (2) Women in Recovery (WiR). All iMUDs met criteria for a DSM-5 diagnosis of Current Amphetamine Use Disorder due to use of Methamphetamine, which was assessed by clinical interview (Mini International Neuropsychiatric Interview 7^32^). Due to high rates of comorbidity, iMUDs were not excluded based on the presence of other substance use disorders or depression/anxiety disorders (for a list of comorbid disorders in the MUD sample, see **Supplemental Table S1**). However, individuals with bipolar disorder, personality disorders, eating disorders, schizophrenia, or obsessive-compulsive disorder were excluded. Current use of psychotropic medications was permitted for iMUDs, as these are frequently utilized by providers in acute substance use treatment. HCs did not have any history of psychiatric illness and were not on any psychotropic medication.

### Protocol

After providing informed consent to participate in a larger study protocol approved by WCG IRB (#20211403), participants completed a drug test and breathalyzer assessment to confirm eligibility for the study. Next, iMUDs completed the Desire for Speed (Methamphetamine) Questionnaire (DSQ)^33^ to assess baseline craving levels.

Following completion of these questionnaires, participants were fit with a silicon mask (see **Figure 1**), which would later be used for anxiety induction during performance of the Horizon Task (described below). This breathing-based anxiety induction apparatus has been used safely and effectively in several previous studies^34–37^. Here, filters are used to add inspiratory resistance (i.e., requiring more effort to breathe in, but no added effort to breathe out), which induces a sensation of air hunger and elevates somatic anxiety. This initial fitting period was part of a sensitivity protocol designed to confirm sufficient comfort with the mask and allow us to assess how anxiety changed as a function of resistance level. During the preliminary sensitivity protocol, participants breathed through the mask while being exposed to six levels of resistance (0, 10, 20, 40, 60, and 80 cmH2O/L/sec) in ascending order for one minute each, with a short break in between each. After each exposure, they were instructed: “Please rate how much anxiety you felt while breathing from 0 to 10” (where 0 indicates no anxiety and 10 indicates maximum possible anxiety). These are referred to as *self-reported anxiety scores*. After completing this protocol, participants removed the mask, and iMUDs completed the DSQ a second time to assess whether craving levels had changed due to anxiety induction.

After this sensitivity protocol, participants completed neuropsychological testing and additional self-report questionnaires as part of the larger study protocol. This ensured participants were able to return to baseline arousal state before performing the Horizon Task. Participants were then re-fit with the mask before task performance and indicated their baseline level of anxiety (using both the self-report item mentioned above and the STAI State scale^38^). Next, they completed two runs of the Horizon Task, where one of the runs included a breathing resistance of 40 cmH2O/L/sec (counterbalanced order across participants). After each run, they again completed the STAI State scale and indicated their self-reported anxiety during task performance.

### Horizon Task

As in previous studies^10^, the Horizon Task here consisted of 80 games in which participants chose between two slot machines with different (unknown) average payout values (see **Figure 1** for a depiction of the task). For one of the slot machines, results were sampled from a Gaussian distribution with a mean of either 40 or 60 and a fixed standard deviation of 8. For the other slot machine, the distribution was shifted 4, 8, 12, 20, or 30 points in either direction from the first slot machine.

Participants first observed outcomes of four forced choices before they were allowed to make either one or six free choices between options to maximize the total number of points received. Games with one or six free choices are referred to as Horizon 1 (H1) and Horizon 6 (H6) games, respectively.

The forced choices in each game were either equally informative (two forced choices for each slot machine) or unequally informative (three forced choices for one slot machine and one for the other). The different information conditions, decision horizons, and mean slot machine values were all counterbalanced throughout the task.

To minimize potential influences on individual differences in behavior, the observed outcome for each choice was sampled from the underlying Gaussian distributions but fixed across participants and task runs. Thus, two participants who chose the same option on a specific trial always observed the same result. However, after preliminary checking of data in the first five participants (all HCs), unexpected behavior in certain games led us to realize that forced choice outcomes in a few cases were not representative of the underlying distributions, which generated concerns given the number of trials per task condition (i.e., with respect to generative mean differences). To minimize this issue, forced choice results in these cases were re-sampled until they more closely aligned with the true differences between underlying distributions. Any potential effects of task version on behavior were accounted for in subsequent analyses.

#### Computational Model

An established computational model was fit to task behavior (i.e., predicting the first free choice across games), as described in detail by Zajkowski, et al. ^39^. In brief, the probability of choosing the right option was calculated using a logistic choice function that included the difference in expected reward values between options, *ΔR*, the information difference between options, *ΔI*, a potential bias toward the right vs. left choice, *B*, and decision noise, *σ*, as follows:

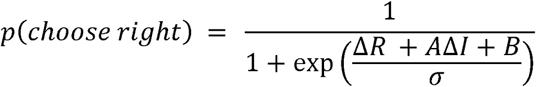

The information difference (*ΔI*) was equal to +1 when one outcome was shown for the right option, –1 when three outcomes were shown for that option, and 0 when two outcomes were shown for each option. This was then scaled by a free parameter referred to as the *information bonus* (depicted above as *A*). The expected reward value difference (*ΔR*) was calculated using a Rescorla-Wagner update equation, where the expected reward value *R* for each option *i* on time step *t* updated based on the prediction error between the expected reward *R_t_^i^* and observed reward *r_i_*. The learning rate *α* varied as a function of uncertainty (i.e., in relation to the number of previous observations):

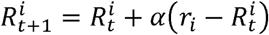

The initial learning rate *α*_0_ was a free parameter fit to participant data. For each subsequent choice, the learning rate was updated with the following equation:

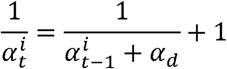

The drift term, *α_d_*, influences how learning rate changes over time. It can also be related to an asymptotic learning rate (*α*_∞_) with the following equation:

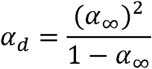

The *α*_∞_ term (bound between 0 and 1) is also a free parameter fit to the data (entailing a value for *α_d_*).

This asymptotic learning rate is the value to which learning rate would theoretically converge if the game were played indefinitely (i.e., due to evolving levels of uncertainty after seeing an increasing number of outcomes). Within the Kalman filter model, *α_d_* reflects the ratio between expected instability (drift) in the underlying reward mean and expected outcome noise around that reward mean. Based on the final equation above, it therefore follows that slower asymptotic learning rates can be seen to reflect an implicit belief that underlying reward means are stable (i.e., minimal drift within a game) and/or that each observed outcome is unreliable (i.e., high levels of outcome noise).

To get parameter estimates for each participant, a hierarchical Bayesian model^40^ with 12 free parameters in total was fit using a Markov Chain Monte Carlo (MCMC) method implemented with MATJAGS^41^. The spatial bias (*B*) and decision noise (*σ*) were fit separately for the four combinations of horizon (H1 or H6) and information condition (equal or unequal). The information bonus (*A*) was fit separately for the two horizon conditions (i.e., this can only be fit for unequal information games). The initial learning rate and asymptotic learning rate were fit across all games together. In our fitting procedure, second-level hyperparameters defined the prior distributions from which individual parameters were sampled (see **Supplemental Table S6** for complete specification of these prior distributions). All participants in both conditions were included under a single second-level prior so that the hyperpriors for HCs and iMUDs were equivalent. This was done to prevent any artificial bias toward group or condition differences.

Previous work has shown that estimates of initial reward expectations trade off with information bonus estimates on this task^39^, as optimistic reward expectations can also promote the choice of unfamiliar options. Thus, given our focus on directed exploration, we fixed the initial reward expectation to a neutral value of 50 to ensure reliable parameter estimates for the information bonuses. We assessed the recoverability of the model by measuring the correlation between parameters used to simulate data and parameters estimated from fitting that simulated data (see **Supplemental Materials** for details).

### Measures

Participants completed the following measures to assess relevant clinical symptoms as well as trait and state psychological characteristics.

### Symptom Severity

To measure MUD severity, we used the Drug Abuse Screening Test (DAST), the Methamphetamine Withdrawal Questionnaire (MAWQ), and the Desires for Speed (Methamphetamine) Questionnaire (DSQ). DAST measures overall drug abuse severity and interference with life functioning^42^; MAWQ measures withdrawal symptoms^43^; and DSQ measures current craving levels^33^. These measures were only gathered in iMUDs.

To measure comorbid symptom dimensions associated with MUD, we used questionnaires measuring depression, anxiety, and impulsivity. Overall depressive symptoms were measured using the Patient Health Questionnaire (PHQ-9)^44^. State and trait anxiety was measured with the State-Trait Anxiety Inventory (STAI-State/Trait)^38,45^. Impulsivity was measured with the Urgency-Premeditation-Perseverance-Sensation Seeking-Positive Urgency (UPPS-P) Impulsive Behavior Scale Total Score^46,47^.

### Cognitive Reflectiveness

The Cognitive Reflection Test [CRT-7^48^] measures the tendency to “stop and think” before immediately trusting one’s intuition. The test asks seven short questions designed such that there is an immediately intuitive, but incorrect, answer, and a correct answer that, while not logically difficult, requires the individual to devote effortful cognitive resources instead of immediately choosing the intuitively appealing response. An example item is “If it takes 5 machines 5 minutes to make 5 widgets, how long would it take 100 machines to make 100 widgets?” (intuitive incorrect answer: 100 minutes; correct answer: 5 minutes).

### Working Memory

The List Sorting Working Memory Test from the NIH Toolbox Cognition Battery^49^ was used to assess working memory. In our analyses, we used participants’ t-scores adjusted for age and sex.

### Statistical Analyses

Between-subject statistical analyses were carried out in R (version 4.4.1) with R Studio. K-means clustering was performed for *α* using the *kmeans* function of the *stats* package^50^. Linear mixed effects models (LMEs) and logistic mixed regressions were run using the *lmer* function and the *glmer* function of the *lme4* package^51^. For the logistic models, the *bobyqa* optimizer was used to estimate the coefficients. In all mixed-effects models, a term for participant ID was included to allow for random intercepts. Effect sizes were calculated with the *F_to_eta2* function of the *effectsize* package^52^. All continuous predictors were mean-centered using the *gscale* function of the *jtools* package^53^. Unless otherwise stated, categorical variables were sum-coded as factors, including group (HCs=–1, iMUDs=1), breathing resistance (absent*=*–1, present*=*1), information condition (equal=–1, unequal=1), horizon (H1=–1, H6=1), sex (male=–1, female=1), and task version (main version=–1, initial version in first 5 participants=1).

The variables age and sex were included in all models as potential covariates to ensure they did not explain observed effects. As a small number of participants also completed a slightly different version of the task (i.e., with a different sequence of reward values sampled from the underlying generative means; described above), task version was also included in all models as a potential covariate. After controlling for age, sex, and task version, a follow-up model was run that additionally included working memory capacity, given that general cognitive ability has previously been shown to positively correlate with performance in the Horizon Task^10^. As working memory data were missing for a subset of participants (N=4), its potential explanatory power was only assessed in the subset of participants with available data in these follow-up analyses (as this would otherwise effectively remove data from these four participants from all analyses). When necessary, significant effects were further interpreted using post-hoc contrasts of estimated marginal trends (EMTs) or estimated marginal means (EMMs) using the *emmeans* package^54^.

### Protocol Validation

To test whether administration of the moderate breathing resistance level (40 cmH2O/L/sec) used during task performance also successfully increased anxiety, an LME was run predicting self-reported anxiety during the task, with resistance condition (baseline, task run with resistance, task run without resistance), group, and their interaction as predictors. Identical LMEs were also run using STAI state scores as the outcome variable in place of self-reported anxiety to confirm consistency (see **Supplemental Table S3** for the results of this model).

To confirm the efficacy of the aversive state induction, we performed another LME to test if administration of the breathing resistance successfully induced anxiety within the pre-task exposure protocol. This model specifically assessed whether self-reported anxiety level was predicted by breathing resistance level (0, 10, 20, 40, 60, and 80 cmH2O/L/sec), group, and/or their interaction (see **Supplemental Table S4** for the results of this model).

### Computational Analysis

Computational measures included: directed exploration (DE), random exploration (RE), initial learning rate (*α*_0_), and asymptotic learning rate (*α*_∞_). DE was calculated by subtracting the information bonus parameter fit to H1 trials from that fit to H6 trials. This allowed us to measure the degree to which participants became more information-seeking as decision horizon increased (i.e., when information became goal-relevant). Note that this only applied to trials in which unequal information was given. RE was calculated by subtracting the decision noise parameter fit to H1 trials from that fit to H6 trials, allowing us to measure the degree to which participants became less value sensitive in their initial choice as decision horizon increased (i.e., which can also serve as an information-seeking strategy). Analyses of RE were here restricted to trials where equal information was given, such that directed information-seeking could not account for any apparent changes in value sensitivity. For analyses with *α*_∞_ as an outcome variable, we also included *α*_0_ as a covariate, given that those with the highest initial learning rate tended to experience the greatest decrease in learning rate over time (somewhat analogous to regression to the mean). For these variables, potential outliers were identified using an iterative Grubb’s method (threshold: *p*<.01), implemented with the *grubbs.test* function from the *outliers* package^55^.

To examine potential effects of group and breathing resistance on each of these model parameters, separate LMEs (and logistic mixed regression for *α*_0_) were run predicting each parameter value from group, resistance condition, and the interaction of those variables. If extreme values of model parameters were identified (testing within each resistance condition separately), analyses were repeated with those data removed, and any discrepancies between results with/without outliers were reported.

To assess whether observed differences in DE and RE were better explained by differences in H1 or H6, separate models were also run with information bonus or decision noise as the outcome variable, respectively, including horizon as an additional predictor. Note that the strength of the interaction between horizon and group can here also be seen as a test of group differences in DE or RE. As a complementary approach, Bayesian analyses were also run using the *brm* function within the *brms* package^56^. These models had the same structure as our frequentist analysis, but additionally incorporated the posterior variance of each parameter estimate for each participant. Bayesian Credible Intervals (BCIs) and Bayes Factors (BFs) were used to compare the likelihood of a model including the interaction between horizon and group to a model omitting that interaction. For the learning rate parameters, this approach was also repeated by comparing models with vs. without an effect of group to assess the strength of evidence for these effects (i.e., consistent with the group difference found for learning rates using the frequentist approach).

To better interpret our main results, we also carried out a test of group differences in avoidance of uncertainty by calculating the (frequency-based) probability of choosing the option with the greater observed mean when it was also the option with greater uncertainty (i.e., with only one forced-choice outcome shown). This was done for the first free choice on H1 and H6 games separately. We then tested an LME predicting this probability for H6 games based on group and resistance condition. The corresponding probability for H1 games was also included in the model to account for any baseline tendency to approach or avoid uncertainty when exploration would not be helpful. An analogous LME was also run to test if negative outcome avoidance differed between groups. This model instead predicted the probability of choosing the option with the greater observed reward mean when it was also the low-uncertainty option (i.e., three observed forced-choice outcomes) in H6 games based on group and resistance condition, while again controlling for baseline tendencies in H1. Here, we reasoned that those with greater negative outcome avoidance would more often choose the option with the higher observed value, despite the greater information gain afforded by the other option.

Finally, we tested if patterns of behavior captured by the model were related to individual differences in somatic anxiety, substance use symptoms, and/or measures of psychopathology. To do so, we first ran LMEs predicting each computational parameter value based on self-reported anxiety level during the task. Among other covariates, the effects of group and resistance condition were also included, as well as the interaction between resistance condition and self-reported anxiety. We next tested LMEs within the iMUD group predicting each parameter value based on DAST, DSQ, or MAWQ scores (tested separately), accounting for the effect of breathing resistance (see **Supplemental Tables S8-S10** for additional details and model results). We were similarly interested in examining if the relationship between parameter values and drug-related symptoms differed between resistance conditions, so these interactions were also included in the models. We then tested LMEs within the iMUD group predicting each parameter value based on the presence of each comorbid affective or substance use disorder (present/absent), accounting for any effect of breathing resistance. Analogous models were also run that included continuous measures of psychopathology (i.e., PHQ-9, STAI-Trait, and UPPS-P) as predictors. To determine if medication status (medicated/unmedicated), time since last use of methamphetamine, and/or time since starting treatment might influence model parameters, similar LMEs were run with those variables (separately).

### Descriptive and Computational Behavioral Analyses

To evaluate the effect of experimental condition on task performance, LMEs tested if free-choice accuracy differed by group, information condition, and/or horizon, and whether these effects might be moderated by breathing resistance and information condition. We additionally tested if these variables predicted accuracy across the free choices of the H6 condition. To examine how model parameters influenced subsequent task performance in H6 trials (i.e., to interpret whether some values might be considered more optimal than others), we also examined if accuracy was predicted by each of the model parameters, free choice number (2-6; i.e., excluding the first free choice to which these parameters were directly fit), resistance, and/or group, and whether a given model parameter moderated the improvement in accuracy as choice number increased (see **Supplemental Table S11** and **S12** for full model results).

### Model Parameters Predicting Cognitive Reflectiveness

We also sought to replicate prior results^10^, and extend them to iMUDs, linking computational Horizon Task metrics (and DE in particular) to cognitive reflectiveness (i.e., CRT scores). We therefore tested if model parameters could be predicted by the number of correct answers on this measure (accounting for effects of resistance). Group was included as a covariate to ensure that observed effects were not explained by group differences in cognitive reflectiveness or parameter values (see **Table 1** and **Figure 1**). If CRT scores significantly predicted a given model parameter, we subsequently tested if CRT scores mediated group differences in that parameter. The *mediate* function within the *mediation* package was used to test for these effects using a nonparametric bootstrapping approach that included 5,000 Monte Carlo simulations. Possible effects of breathing resistance were also incorporated into these analyses.

After testing for potential mediation effects, we further determined if any observed relationships were specific to the iMUD population by testing LMEs predicting computational parameter values based on CRT scores.

## Data Availability

All data produced in the present study are available upon reasonable request to the authors.

## Author Contributions

Writing: C.M.G., T.T., and R.S. Review and editing: all authors. Data analysis: C.M.G., T.T., C.A.L., R.S, and R.H. Methodology: R.S and R.C.W. Conceptualization: R.S., J.L.S, R.C.W., S.S.K., M.P.P. Funding acquisition: R.S. and M.P.P. Data collection: A.E.C., S.T., C.A.L., N.L. Project administration: R.S.

## Data and Code Availability

All data and code used in the formulation of this manuscript are included as supplemental files and available at: https://github.com/Smith-Lab-LIBR/Kalman_Filter_Model_Horizon_Task_Methamphetamine_Study.

## Acknowledgements

This project was funded by National Institute of General Medical Sciences (P20GM121312 [R.S. and M.P.P.]) and the Laureate Institute for Brain Research.

