## Supplemental Materials for "Individuals with Methamphetamine Use Disorder Show Reduced Directed Exploration and Learning Rates Independent of an Aversive Interoceptive State Induction"

**Diagnostic Categorization for Individuals with Methamphetamine Use Disorder**

Participants were screened based on the Mini International Neuropsychiatric Interview 7^1^. All individuals with Methamphetamine Use Disorder (iMUDs) met the criteria for a DSM-5 diagnosis of Current Amphetamine Use Disorder due to use of Methamphetamine. The prevalence of other comorbid disorders within the iMUD group is shown in **Table S1**.

**Table S1. Comorbidity Table for Participants with Methamphetamine Use Disorder**

| **Disorder** | **AUD** | **Opioid** | **Cann.** | **Other SUD** | **Sedative** | **Cocaine** | **MDD** | **Anti–social** | **PTSD** | **GAD** | **SAD** |
| --- | --- | --- | --- | --- | --- | --- | --- | --- | --- | --- | --- |
| **AUD** | 18 |  |  |  |  |  |  |  |  |  |  |
| **Opioid** | 5 | 18 |  |  |  |  |  |  |  |  |  |
| **Cann.** | 4 | 1 | 15 |  |  |  |  |  |  |  |  |
| **Other SUD** | 0 | 2 | 0 | 3 |  |  |  |  |  |  |  |
| **Sedative** | 1 | 1 | 0 | 0 | 2 |  |  |  |  |  |  |
| **Cocaine** | 1 | 0 | 2 | 0 | 0 | 2 |  |  |  |  |  |
| **MDD** | 7 | 5 | 3 | 1 | 2 | 0 | 14 |  |  |  |  |
| **AntiSoc.** | 5 | 3 | 4 | 0 | 0 | 0 | 3 | 8 |  |  |  |
| **PTSD** | 0 | 0 | 1 | 0 | 0 | 0 | 1 | 0 | 1 |  |  |
| **GAD** | 1 | 1 | 0 | 0 | 0 | 0 | 1 | 1 | 0 | 1 |  |
| **SAD** | 0 | 0 | 0 | 0 | 1 | 0 | 1 | 0 | 0 | 1 | 1 |

**Note**. All 56 individuals with MUD (iMUDs) received a diagnosis of Amphetamine Use Disorder. Additional diagnoses are listed in the table above, where AUD = Alcohol Use Disorder, Opioid = Opioid Use Disorder, Cann. = Cannabis Use Disorder, Other SUD = Other Substance Use Disorder, Sedative = Sedative, Hypnotic, or Anxiolytic Use Disorder, Cocaine = Cocaine Use Disorder, MDD = Major Depressive Disorder, Antisocial = Antisocial Personality Disorder, PTSD = Posttraumatic Stress Disorder, GAD = Generalized Anxiety Disorder, SAD = Social Anxiety Disorder.

***Anxiety During the Task***

Results for the linear mixed-effect models (LMEs) predicting self-reported anxiety based on group, resistance condition (baseline, task run without resistance, and task run with resistance), and their interaction are shown in **Table S2**. Age, sex, task version, and working memory were included in the model as covariates. Coefficients were estimated with reference to baseline anxiety. Results of an analogous model predicting STAI State Anxiety are shown in **Table S3**.

**Table S2 LME Predicting Self-Reported Anxiety During the Task**

| **Predictor** | **Statistical Results** |
| --- | --- |
| **Group** | *F*(1, 103.0)=17.86, ***p<*.001**, $\eta_{p}^{2}$=0.15, *b=*0.69 [-0.098, 1.477] |
| **Resistance Condition** | *F*(2, 214.0)=83.65, ***p<*.001**, $\eta_{p}^{2}$=0.44,  *b*(task run without resistance)=0.14 [-0.337, 0.623],  *b*(task run with resistance)=1.38 [0.895, 1.855] |
| **Interaction** | *F*(2, 214.0)=10.75, ***p<*.001**, $\eta_{p}^{2}$=0.09,  *b*(group effect \| task run without resistance)=0.71 [0.017, 1.395],  *b*(group effect \| task run with resistance)=1.63 [0.936, 2.314] |

**Table S3. LME Predicting STAI Anxiety During the Task**

| **Predictor** | **Statistical Results** |
| --- | --- |
| **Group** | *F*(1, 103.0)=8.67, ***p=*.004**, $\eta_{p}^{2}$=0.08, *b=*2.86 [-1.450, 7.161] |
| **Resistance Condition** | *F*(2, 214.0)=115.14, ***p<*.001**, $\eta_{p}^{2}$=0.52,  *b*(task run without resistance)=1.91 [-0.273, 4.094],  *b*(task run with resistance)=9.20 [7.013, 11.380] |
| **Interaction** | *F*(2, 214.0)=5.98, ***p=*.003**, $\eta_{p}^{2}$=0.05  *b*(group effect \| task run without resistance)=3.62 [0.487, 6.749],  *b*(group effect \| task run with resistance)=5.43 [2.295, 8.557] |

***Anxiety During the Resistance Sensitivity Protocol***

Results for the LMEs predicting self-reported anxiety during the resistance sensitivity protocol by group, continuous resistance level (0, 10, 20, 40, 60, and 80 cmH2O/L/sec), and their interaction are shown in **Table S4**. Age, sex, task version, and working memory were included in the model as covariates.

**Table S4. LME Predicting Self-Reported Anxiety During the Resistance Sensitivity Protocol**

| **Predictor** | **Statistical Results** |
| --- | --- |
| **Group** | *F*(1, 103.1)=8.49, ***p=.*004**, $\eta_{p}^{2}$=0.076, *b=*0.658 [0.219, 1.098] |
| **Resistance Level** | *F*(1, 539.5)=364.75, ***p*<.001**, $\eta_{p}^{2}$=0.403, *b=*0.041 [0.036, 0.045] |
| **Interaction** | *F*(1, 539.5)=23.83**, *p*<.001**, $\eta_{p}^{2}$=0.042, *b=*0.01 [0.006, 0.015] |

***Task Performance***

We tested an LME predicting first free-choice accuracy in the Horizon Task based on group, information condition (equal/unequal information games), horizon (H1/H6), breathing resistance (present/absent), and the three-way interactions of horizon, group, and breathing resistance, as well as horizon, group, and information condition (including respective two-way interactions; see **Table S5**). Age, sex, task-version, and working memory were included in the model as covariates. Post-hoc analysis showed that the significant interaction between horizon and group was due to HCs having a greater increase in accuracy from H6 to H1 (EMM=.09) than iMUDs (EMM=.05).

**Table S5. LME Predicting First Free Choice Accuracy in the Horizon Task**

| **Predictor** | **Statistical Results** |
| --- | --- |
| **Horizon** | *F*(1, 753.0)=131.99, ***p*<.001**, $\eta_{p}^{2}$=0.15, *b=*-0.037 [-0.043, -0.031] |
| **Group** | *F*(1, 103.0)=20.91, ***p*<.001**, $\eta_{p}^{2}$=0.17, *b=*-0.062 [-0.088, -0.035] |
| **Info Condition** | *F*(1, 753.0)=74.32, ***p*<.001**, $\eta_{p}^{2}$=0.09, *b=*-0.028 [-0.034, -0.022] |
| **Resistance** | *F*(1, 753.0)=0.00, *p=.*958, $\eta_{p}^{2}$<0.01, *b=*0.00 [-0.006, 0.006] |
| **Horizon x Group** | *F*(1, 753.0)=8.76, ***p=.*003**, $\eta_{p}^{2}$=0.01, *b=*0.01 [0.003, 0.016] |
| **Horizon x Info Condition** | *F*(1, 753.0)=0.88, *p=.*348, $\eta_{p}^{2}$<0.01, *b=*0.003 [-0.003, 0.009] |
| **Group x Info Condition** | *F*(1, 753.0)=0.46, *p=.*497, $\eta_{p}^{2}$<0.01, *b=*-0.002 [-0.009, 0.004] |
| **Group x Resistance** | *F*(1, 753.0)=0.16, *p=.*685, $\eta_{p}^{2}$<0.01, *b=*-0.001 [-0.008, 0.005] |
| **Horizon x Resistance** | *F*(1, 753.0)=0.03, *p=.*873, $\eta_{p}^{2}$<0.01, *b=-*0.0006 [-0.007, 0.006] |
| **Horizon x Group x Info Condition** | *F*(1, 753.0)=0.40, *p*=.529, $\eta_{p}^{2}$<0.01, *b=*-0.002 [-0.008, 0.004] |
| **Horizon x Group x Resistance** | *F*(1, 753.0)=0.824, *p=.*364, $\eta_{p}^{2}$<0.01, *b=*-0.003 [-0.009, 0.003] |

***Testing if Parameter Values Differed by Group or Resistance Condition***

A hierarchical Bayesian model^2^ was fit using a Markov Chain Monte Carlo (MCMC) method implemented with MATJAGS^3^. The specification of the prior distributions for each free parameter is listed in **Table S6**. The correlations between each estimated free parameter and the other parameters in the model are shown in **Figure S1**. **Figure S2** shows the recoverability for each parameter in the model, assessed by the correlation between parameters used to simulate data and parameters estimated from the simulated data. For the recoverability analyses, the parameters estimated from participants’ actual data (N=114) were used to simulate plausible behavior under the model.

**Table S6. Computational Model Parameters, Priors, Hyperparameters, and Hyperpriors**

| **Parameter** | **Prior** | **Hyperparameters** | **Hyperpriors** |
| --- | --- | --- | --- |
| Initial Learning Rate $\alpha_{0}$ | $\alpha_{0} \sim Beta(a_{a_{0}}{,b}_{a_{0}})$ | $\theta_{\alpha_{0}}=Beta(a_{a_{0}}{,b}_{a_{0}})$ | $a_{a_{0}}\sim Uniform\left( 0.1,10 \right)$  $b_{a_{0}}\sim Uniform\left( 0.5,10 \right)$ |
| Asymptotic Learning Rate $\alpha_{\infty}$ | $\alpha_{\infty} \sim Beta(a_{\alpha_{\infty}}{,b}_{\alpha_{\infty}})$ | $\theta_{\alpha_{\infty}}=Beta(a_{\alpha_{\infty}}{,b}_{\alpha_{\infty}})$ | $a_{\alpha_{\infty}}\sim Uniform\left( 0.1,10 \right)$  $b_{\alpha_{\infty}}\sim Uniform\left( 0.5,10 \right)$ |
| Information Bonus $A$ | $A \sim Gaussian(\mu_{A},\sigma_{A}$) | $\theta_{A}=Gaussian(\mu_{A},\sigma_{A}$) | $\mu_{A} \sim Gaussian\left( 0,100 \right)$  $\sigma_{A} \sim Gamma\left( 1,0.001 \right)$ |
| Spatial Bias $B$ | $B \sim Gaussian(\mu_{B},\sigma_{B}$) | $\theta_{B}=Gaussian(\mu_{B},\sigma_{B}$) | $\mu_{B} \sim Gaussian\left( 0,100 \right)$  $\sigma_{B} \sim Gamma\left( 1,0.001 \right)$ |
| Decision Noise $\sigma$ | $\sigma\sim Gamma(k_{\sigma},\lambda_{\sigma})$ | $\theta_{\sigma}= Gamma(k_{\sigma},\lambda_{\sigma})$ | $k_{\sigma} \sim Exp(0.1)$  $\lambda_{\sigma} \sim Exp(10)$ |

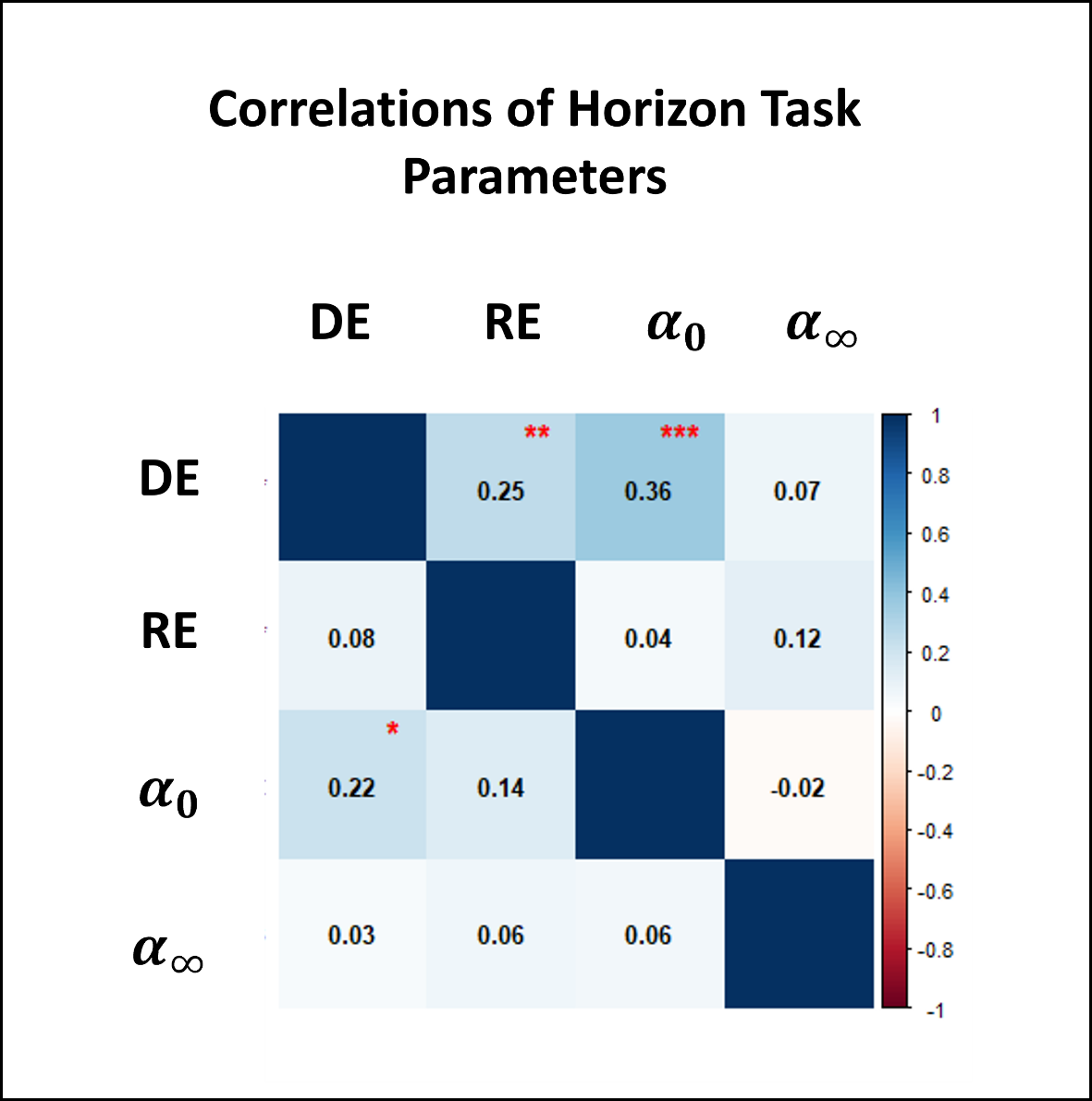

**Figure S1. Correlations of Horizon Task Parameters.** Note DE=Directed Exploration, RE=Random Exploration, $\alpha_{0}$=Initial Learning Rate, $\alpha_{\infty}$=Asymptotic Learning Rate controlling for Initial Learning Rate (comprising the residuals of the linear model predicting $\alpha_{\infty}$ based on $\alpha_{0}$ separately for each resistance condition). Correlations above the diagonal show the relationships between parameters for the task run without breathing resistance; correlations below the diagonal show relationships for the task run with breathing resistance. Note that Spearman’s rank correlation was used for $\alpha_{0}$ given its bimodality.

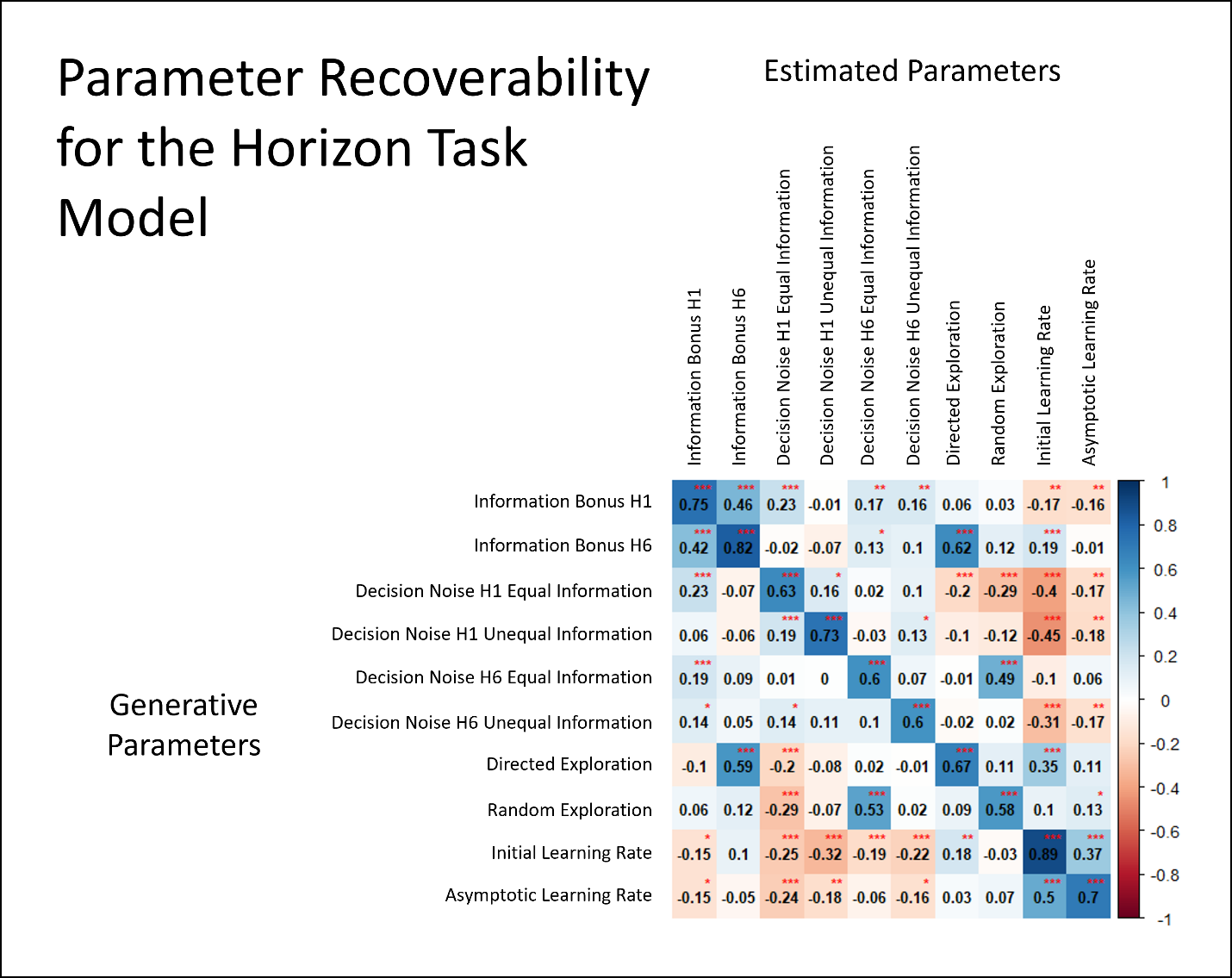
**Figure S2. Parameter Recoverability for the Horizon Task Model.** The parameters used to simulate data are shown on the left as *Generative Parameters*; the parameters estimated from the simulated data are shown above as *Estimated Parameters*. The strong correlations between generative and estimated parameters (*r*=.60 to *r*=.89) indicate good recoverability.

Results for the LMEs (and logistic mixed-effects models for $\alpha_{0}$) predicting model parameters based on group, resistance, and their interaction are shown in **Table S7**. Age, sex, task version, and working memory were included in each model as covariates.

**Table S7. Results for Models Predicting Parameter Values Based on Group and Resistance Including Working Memory as a Covariate**

|  | **DE** | **RE** | $\boldsymbol{\alpha}_{\boldsymbol{0}}$ | $\boldsymbol{\alpha}_{\boldsymbol{\infty}}$ |
| --- | --- | --- | --- | --- |
| **Group** | *F*(1,103.0)=3.09,  *p*=.082,  $\eta_{p}^{2}$=.03,  *b*=-0.614 [-1.298, 0.071] | *F*(1,103.0)=4.60,  ***p*=.034**,  $\eta_{p}^{2}$=.04,  *b*=-0.377 [-0.721, -0.033] | $\chi^{2}$(1)=8.60,  ***p*=.003**,  *b*=-1.419 [-2.363, -0.476] | *F*(1,109.2)=7.95,  ***p*=.006**,  $\eta_{p}^{2}$=.07,  *b*=-0.038 [-0.064, -0.011] |
| **Resistance** | *F*(1,107.0)=2.17,  *p*=.144,  $\eta_{p}^{2}$=.02,  *b*=-0.221 [-0.523, 0.081] | *F*(1,107.0)=0.26,  *p*=.612,  $\eta_{p}^{2}$<.01,  *b*=-0.054 [-0.281, 0.172] | $\chi^{2}$(1)=0.15,  *p*=.695,  *b*=-0.081 [-0.468, 0.307] | *F*(1,106.5)=1.76,  *p*=.187,  $\eta_{p}^{2}$=.02,  *b*=0.009 [-0.004, 0.022] |
| **Group x Resistance** | *F*(1,107.0)=1.60,  *p*=.208,  $\eta_{p}^{2}$=.01,  *b*=0.195 [-0.107, 0.497] | *F*(1,107.0)=1.79,  *p*=.184,  $\eta_{p}^{2}$=.02,  *b*=0.154 [-0.072, 0.380] | $\chi^{2}$(1)=0.17,  *p*=.683,  *b*=-0.081 [-0.468, 0.307] | *F*(1,106.9)=0.43,  *p*=.513,  $\eta_{p}^{2}$<.01,  *b*=0.004 [-0.009, 0.017] |

***Model Parameters and Substance Use Symptoms***

We tested if each model parameter could be predicted by DAST and MAWQ (separately). In these models, we included the interaction between resistance and symptom measure (and main effect of resistance) to test if the relationship between each model parameter and symptom severity differed by task condition (resistance present/absent). An additional set of models tested if each parameter could be predicted by the change in DSQ from somatic anxiety induction and baseline DSQ (see **Table S10**). To ensure that the effects of somatic anxiety induction were not accounted for by baseline DSQ differences, we instead used the residuals derived from a linear regression using baseline DSQ as a predictor of the change in DSQ scores (referred to as DSQ-Change). In these models, we likewise included the interaction between resistance and DSQ-Change as well as the interaction between resistance and baseline DSQ (and potential main effects) to test if breathing resistance moderated the effect of either variable. Unfortunately, these models had less statistical power given that only 39 iMUDs completed the DSQ before and after anxiety induction, because the measure was added part-way into the study. Age, sex, and working memory were included in all models as covariates (task version was not included, as all iMUDs completed the same version of the task). Given that one participant had a DSQ-Change value that was an outlier (on the high end) based on a Grubb’s test, their data were excluded from models testing this effect.

In all models, we did not observe a significant relationship between substance use symptoms and computational parameters, nor did we observe an interaction effect between substance use symptoms and breathing resistance (see **Tables S8-S10**). Surprisingly, in the LME predicting $\alpha_{\infty}$ that included MAWQ as a predictor, there was an effect of breathing resistance such that $\alpha_{\infty}$ was higher for the task run with breathing resistance (EMM=.24) than without breathing resistance (EMM=.21; see **Table S8**). In an analogous model that instead included DAST as a predictor, we observed a similar effect of breathing resistance (**Table S9**). However, we treat these results with caution considering the absence of an effect of breathing resistance in all other analyses with computational parameters.

**Table S8. Model Parameters and MAWQ**

|  | **DE** | **RE** | $\boldsymbol{\alpha}_{\boldsymbol{0}}$ | $\boldsymbol{\alpha}_{\boldsymbol{\infty}}$ |
| --- | --- | --- | --- | --- |
| **MAWQ** | F(1,48.0)=0.08,  p=.781,  $\eta_{p}^{2}$<.01,  b=-0.016 [-0.126, 0.095] | *F*(1,48.0)=0.25,  *p*=.616,  $\eta_{p}^{2}$<.01,  *b*=0.015 [-0.043, 0.073] | *F*(1,48.0)=0.92,  *p*=.342,  $\eta_{p}^{2}$=.02,  *b*=-0.006 [-0.018, 0.006] | *F*(1,47.5)=0.00,  *p*=.978,  $\eta_{p}^{2}$<.01,  *b*=0.000 [-0.005, 0.005] |
| **Resistance** | *F*(1,51.0)=0.02,  *p*=.901,  $\eta_{p}^{2}$<.01,  *b*=-0.022 [-0.437, 0.392] | *F*(1,51.0)=0.56,  *p*=.457,  $\eta_{p}^{2}$=.01,  *b*=0.092 [-0.169, 0.354] | *F*(1,51.0)=1.73,  *p*=.194,  $\eta_{p}^{2}$=.03,  *b*=-0.017 [-0.042, 0.008] | *F*(1,51.2)=4.81,  ***p*=.033**,  $\eta_{p}^{2}$=.09,  *b*=0.015 [0.002, 0.029] |
| **MAWQ x Resistance** | *F*(1,51.0)=0.33,  *p*=.568,  $\eta_{p}^{2}$<.01,  *b*=0.018 [-0.043, 0.078] | *F*(1,51.0)=2.66,  *p*=.109,  $\eta_{p}^{2}$=.05,  *b*=-0.032 [-0.070, 0.006] | *F*(1,51.0)=0.06,  *p*=.812,  $\eta_{p}^{2}$<.01,  *b*=-0.000 [-0.004, 0.003] | *F*(1,50.4)=0.23,  *p*=.635,  $\eta_{p}^{2}$<.01,  *b*=0.000 [-0.001, 0.002] |

**Table S9. Model Parameters and DAST**

|  | **DE** | **RE** | $\boldsymbol{\alpha}_{\boldsymbol{0}}$ | $\boldsymbol{\alpha}_{\boldsymbol{\infty}}$ |
| --- | --- | --- | --- | --- |
| **DAST** | *F*(1,48.0)=1.02,  *p*=.318,  $\eta_{p}^{2}$=.02,  *b*=0.199 [-0.187, 0.586] | *F*(1,48.0)=2.63,  *p*=.111,  $\eta_{p}^{2}$=.05,  *b*=0.166 [-0.035, 0.367] | *F*(1,48.0)=2.44,  *p*=.125,  $\eta_{p}^{2}$=.05,  *b*=0.034 [-0.009, 0.076] | *F*(1,48.2)=1.48,  *p*=.230,  $\eta_{p}^{2}$=.03,  *b*=0.010 [-0.006, 0.027] |
| **Resistance** | *F*(1,51.0)=0.02,  *p*=.902,  $\eta_{p}^{2}$<.01,  *b*=-0.027 [-0.442, 0.389] | *F*(1,51.0)=0.54,  *p*=.467,  $\eta_{p}^{2}$=.01,  *b*=0.101 [-0.167, 0.368] | *F*(1,51.0)=1.73,  *p*=.194,  $\eta_{p}^{2}$=.03,  *b*=-0.017 [-0.042, 0.008] | *F*(1,51.2)=4.82,  ***p*=.033**,  $\eta_{p}^{2}$=.09,  *b*=0.015 [0.002, 0.028] |
| **DAST x Resistance** | *F*(1,51.0)=0.06,  *p*=.807,  $\eta_{p}^{2}$<.01,  *b*=-0.027 [-0.246, 0.191] | *F*(1,51.0)=0.30,  *p*=.586,  $\eta_{p}^{2}$<.01,  *b*=0.039 [-0.101, 0.180] | *F*(1,51.0)=0.03,  *p*=.869,  $\eta_{p}^{2}$<.01,  *b*=-0.001 [-0.014, 0.012] | *F*(1,50.4)=0.83,  *p*=.367,  $\eta_{p}^{2}$=.02,  *b*=-0.003 [-0.010, 0.004] |

**Table S10. Model Parameters and DSQ Before and After Somatic Anxiety Induction**

|  | **DE** | **RE** | $\boldsymbol{\alpha}_{\boldsymbol{0}}$ | $\boldsymbol{\alpha}_{\boldsymbol{\infty}}$ |
| --- | --- | --- | --- | --- |
| **DSQ-Change** | *F*(1,31.0)=0.30,  *p*=.587,  $\eta_{p}^{2}$<.01,  *b*=-0.019 [-0.086, 0.048] | *F*(1,31.0)=1.50,  *p*=.230,  $\eta_{p}^{2}$=.05,  *b*=0.022 [-0.013, 0.058] | *F*(1,31.0)=0.02,  *p*=.882,  $\eta_{p}^{2}$<.01,  *b*=-0.001 [-0.007, 0.006] | *F*(1,30.1)=0.73,  *p*=.399,  $\eta_{p}^{2}$=.02,  *b*=-0.001 [-0.004, 0.001] |
| **Baseline DSQ** | *F*(1,31.0)=0.06,  *p*=.814,  $\eta_{p}^{2}$<.01,  *b*=-0.003 [-0.031, 0.024] | *F*(1,31.0)=0.00,  *p*=.983,  $\eta_{p}^{2}$<.01,  *b*=-0.000 [-0.015, 0.014] | *F*(1,31.0)=0.15,  *p*=.704,  $\eta_{p}^{2}$<.01,  *b*=0.001 [-0.002, 0.003] | *F*(1,30.2)=0.03,  *p*=.865,  $\eta_{p}^{2}$<.01,  *b*=0.000 [-0.001, 0.001] |
| **Resistance** | *F*(1,34.0)=0.63,  *p*=.431,  $\eta_{p}^{2}$=.02,  *b*=-0.214 [-0.722, 0.294] | *F*(1,34.0)=0.05,  *p*=.829,  $\eta_{p}^{2}$<.01,  *b*=0.036 [-0.277, 0.349] | *F*(1,34.0)=1.29,  *p*=.265,  $\eta_{p}^{2}$=.04,  *b*=-0.016 [-0.045, 0.014] | *F*(1,34.0)=1.13,  *p*=.296,  $\eta_{p}^{2}$=.03,  *b*=0.010 [-0.008, 0.027] |
| **Resistance x DSQ-Change** | *F*(1,34.0)=0.13,  *p*=.718,  $\eta_{p}^{2}$<.01,  *b*=-0.007 [-0.044, 0.030] | *F*(1,34.0)=0.57,  *p*=.455,  $\eta_{p}^{2}$=.02,  *b*=-0.009 [-0.032, 0.014] | *F*(1,34.0)=0.92,  *p*=.344,  $\eta_{p}^{2}$=.03,  *b*=-0.001 [-0.003, 0.001] | *F*(1,33.9)=0.61,  *p*=.439,  $\eta_{p}^{2}$=.02,  *b*=0.001 [-0.001, 0.002] |
| **Resistance x Baseline DSQ** | *F*(1,34.0)=0.65,  *p*=.427,  $\eta_{p}^{2}$=.02,  *b*=0.006 [-0.009, 0.021] | *F*(1,34.0)=0.00,  *p*=.971,  $\eta_{p}^{2}$<.01,  *b*=0.000 [-0.009, 0.009] | *F*(1,34.0)=3.17,  *p*=.084,  $\eta_{p}^{2}$=.09,  *b*=-0.001 [-0.002, 0.000] | *F*(1,34.8)=0.15,  *p*=.703,  $\eta_{p}^{2}$<.01,  *b*=-0.000 [-0.001, 0.000] |

***Relationship Between Task Accuracy and Model Parameters***

We tested LMEs predicting task accuracy based on model parameters in both information conditions separately (**Table S11** and **Table S12**). Separate models were run with information bonus, decision noise, $\alpha_{0}$, or $\alpha_{\infty}$ as a predictor, additionally including main effects of breathing resistance condition, group, free choice number (2–6; i.e., excluding the first free choice to which these parameters were directly fit), and the interaction between the given model parameter and choice number. Age, sex, task version, and working memory were also included as covariates.

**Table S11. Results for Models Predicting Task Accuracy in the Unequal Information Games**

|  | **Information Bonus** | $\boldsymbol{\alpha}_{\boldsymbol{0}}$ | $\boldsymbol{\alpha}_{\boldsymbol{\infty}}$ |
| --- | --- | --- | --- |
| **Parameter** | *F*(1,758.2)=1.72,  *p*=.191,  $\eta_{p}^{2}$<.01,  *b*=0.002 [-0.001, 0.005] | *F*(1,717.6)=52.06,  ***p*<.001**,  $\eta_{p}^{2}$=.07,  *b*=0.139 [0.101, 0.176] | *F*(1,1014.2)=1.67,  *p*=.197,  $\eta_{p}^{2}$<.01,  *b*=0.048 [-0.025, 0.121] |
| **Resistance** | *F*(1,1013.2)=0.09,  *p*=.767,  $\eta_{p}^{2}$<.01,  *b*=0.001 [-0.005, 0.006] | *F*(1,977.8)=0.04,  p=.839,  $\eta_{p}^{2}$<.01,  *b*=0.001 [-0.005, 0.006] | *F*(1,981.5)=0.04,  *p*=.840,  $\eta_{p}^{2}$<.01,  *b*=-0.001 [-0.006, 0.005] |
| **Group** | *F*(1,103.3)=16.53,  ***p*<.001**,  $\eta_{p}^{2}$=.14,  *b*=-0.054 [-0.080, -0.028] | *F*(1,109.7)=11.58,  ***p*<.001**,  $\eta_{p}^{2}$=.10,  *b*=-0.038 [-0.059, -0.016] | *F*(1,106.5)=16.64,  ***p*<.001**,  $\eta_{p}^{2}$=.14,  *b*=-0.053 [-0.078, -0.027] |
| **Choice Number** | *F*(1,977.1)=54.20,  ***p*<.001**,  $\eta_{p}^{2}$=.05,  *b*=0.014 [0.010, 0.017] | *F*(1,977.1)=54.20,  ***p*<.001**,  $\eta_{p}^{2}$=.05,  *b*=0.014 [0.010, 0.017] | *F*(1,977.0)=53.61,  ***p*<.001**,  $\eta_{p}^{2}$=.05,  *b*=0.014 [0.010, 0.017] |
| **Parameter x Choice Number** | *F*(1,977.1)=3.27,  *p*=.071,  $\eta_{p}^{2}$<.01,  *b*=0.001 [-0.000, 0.002] | *F*(1,977.1)=4.94,  ***p*=.026**,  $\eta_{p}^{2}$<.01,  *b*=-0.013 [-0.024, -0.002] | *F*(1,977.0)=0.01,  *p*=.924,  $\eta_{p}^{2}$<.01,  *b*=0.001 [-0.024, 0.026] |

**Table S12. Results for Models Predicting Task Accuracy in the Equal Information Condition Games**

|  | **Decision Noise** | $\boldsymbol{\alpha}_{\boldsymbol{0}}$ | $\boldsymbol{\alpha}_{\boldsymbol{\infty}}$ |
| --- | --- | --- | --- |
| **Parameter** | *F*(1,1076.5)=0.04,  *p*=.840,  $\eta_{p}^{2}$<.01,  *b*=-0.000 [-0.005, 0.004] | *F*(1,804.7)=20.51,  ***p*<.001**,  $\eta_{p}^{2}$=.02,  *b*=0.093 [0.053, 0.133] | *F*(1,1014.2)=6.62,  ***p*=.010**,  $\eta_{p}^{2}$<.01,  *b*=0.099 [0.024, 0.174] |
| **Resistance** | *F*(1,977.1)=0.29,  *p*=.593,  $\eta_{p}^{2}$<.01,  *b*=0.001 [-0.004, 0.007] | *F*(1,977.8)=0.49,  *p*=.483,  $\eta_{p}^{2}$<.01,  *b*=0.002 [-0.003, 0.007] | *F*(1,981.5)=0.06,  *p*=.814,  $\eta_{p}^{2}$<.01,  *b*=0.001 [-0.005, 0.006] |
| **Group** | *F*(1,102.9)=14.95,  ***p*<.001**,  $\eta_{p}^{2}$=.13,  *b*=-0.054 [-0.081, -0.026] | *F*(1,109.2)=11.03,  ***p*=.001**,  $\eta_{p}^{2}$=.09,  *b*=-0.042 [-0.066, -0.017] | *F*(1,106.5)=13.16,  ***p*<.001**,  $\eta_{p}^{2}$=.11,  *b*=-0.049 [-0.075, -0.022] |
| **Choice Number** | *F*(1,977.0)=68.40,  ***p*<.001**,  $\eta_{p}^{2}$=.07,  *b*=0.016 [0.012, 0.020] | *F*(1,977.1)=67.66,  ***p*<.001**,  $\eta_{p}^{2}$=.06,  *b*=0.016 [0.012, 0.020] | *F*(1,977.0)=68.02,  ***p*<.001**,  $\eta_{p}^{2}$=.07,  *b*=0.016 [0.012, 0.020] |
| **Parameter x Choice Number** | *F*(1,977.0)=4.17,  ***p*=.041**,  $\eta_{p}^{2}$<.01,  *b*=0.002 [0.000, 0.004] | *F*(1,977.1)=0.78,  *p*=.379,  $\eta_{p}^{2}$<.01,  *b*=0.005 [-0.007, 0.017] | *F*(1,977.0)=2.62,  *p*=.106,  $\eta_{p}^{2}$<.01,  *b*=0.021 [-0.004, 0.047] |
